# Genetic insights into ossification of the posterior longitudinal ligament of the spine

**DOI:** 10.1101/2022.06.16.22276152

**Authors:** Yoshinao Koike, Masahiko Takahata, Masahiro Nakajima, Nao Otomo, Hiroyuki Suetsugu, Xiaoxi Liu, Tsutomu Endo, Shiro Imagama, Kazuyoshi Kobayashi, Takashi Kaito, Satoshi Kato, Yoshiharu Kawaguchi, Masahiro Kanayama, Hiroaki Sakai, Takashi Tsuji, Takeshi Miyamoto, Hiroyuki Inose, Toshitaka Yoshii, Masafumi Kashii, Hiroaki Nakashima, Kei Ando, Yuki Taniguchi, Kazuhiro Takeuchi, Shuji Ito, Kohei Tomizuka, Keiko Hikino, Yusuke Iwasaki, Yoichiro Kamatani, Shingo Maeda, Hideaki Nakajima, Kanji Mori, Atsushi Seichi, Shunsuke Fujibayashi, Tsukasa Kanchiku, Kei Watanabe, Toshihiro Tanaka, Kazunobu Kida, Sho Kobayashi, Masahito Takahashi, Kei Yamada, Hiroshi Takuwa, Hsing-Fang Lu, Shumpei Niida, Kouichi Ozaki, Yukihide Momozawa, Genetic Study Group of Investigation Committee on Ossification of the Spinal Ligaments, Masashi Yamazaki, Atsushi Okawa, Morio Matsumoto, Norimasa Iwasaki, Chikashi Terao, Shiro Ikegawa

## Abstract

**Background:** Ossification of the posterior longitudinal ligament of the spine (OPLL) is an intractable disease, leading to severe neurological deficits. Its etiology and pathogenesis are mostly unknown. The relationship between OPLL and comorbidities, especially type 2 diabetes (T2D) and body mass index (BMI), has been the focus of attention; however, no trait has been proven to have a causal relationship.

**Methods:** To clarify the etiology and pathogenesis of OPLL, we conducted a meta-analysis of genome-wide association studies (GAWSs) using 22,016 Japanese individuals. We classified OPLL into cervical, thoracic and the other types, and conducted GWAS sub-analyses. We conducted a gene-based association analysis and a transcriptome-wide Mendelian randomization approach to identify other potential causal genes. To investigate cell groups related to OPLL, we conducted cell type group enrichment analysis. To identify traits with a causal effect on OPLL, we evaluated the genetic correlation with 99 complex traits and then performed mendelian randomization (MR) analyses. Finally, we generated polygenic risk score (PRS) to investigate the genetic impact of the causal trait on OPLL subtypes.

**Results:** A GWAS meta-analysis identified 14 significant loci, including eight novel loci. GWAS sub-analyses identified subtype-specific signals. A Gene-based association analysis and a transcriptome-wide Mendelian randomization approach identified five and three potential causal genes, respectively. These loci/genes contained bone metabolism-related genes. Cell type group enrichment analysis observed significant enrichment of the polygenic signals in the active enhancers of the connective/bone cell group, especially H3K27ac in chondrogenic differentiation cells. Genetic correlations showed positive correlation with T2D and BMI and negative correlation with cerebral aneurysm and osteoporosis. MR analyses demonstrated a significant causal effect of increased BMI and high bone mineral density (BMD) on OPLL, but not of T2D, indicating that high BMI confounded the T2D correlation. A PRS for BMI demonstrated that the effect of BMI was particularly strong in thoracic OPLL.

**Conclusion:** We identified multiple causative genes involved in bone metabolism that are candidates for future therapeutic targets. By MR analyses, we showed for the first time a causal relationship between the common metabolic conditions (high BMI and BMD) and OPLL. We successfully linked intervenable traits to OPLL.

## Introduction

Ossification of the posterior longitudinal ligament of the spine (OPLL) is caused by progressive ectopic ossification of the posterior longitudinal ligament of the spine. It can occur at any spine level from the cervical to the lumbar spine, and ossified ligaments compress the spinal cord and roots, leading to severe neurologic deficit (Matsunaga and Sakou, 2012). OPLL is a common disease affecting ∼1.7% of the population, with slight differences among ancestries (∼4.3% in Japan) (Endo et al., 2020; Matsunaga and Sakou, 2012). However, its etiology and pathogenesis remain unknown. Histological studies suggest that OPLL develops through endochondral ossification (Sato et al., 2007; Sugita et al., 2013). At present, there is no therapeutic or preventive measure for OPLL other than surgery to decompress the spinal cord and roots. Therefore, it is necessary to clarify its etiology and pathogenesis to develop effective measures to prevent and treat OPLL.

OPLL is assumed to be a polygenic disease where complex genetic and environmental factors interact. Epidemiological studies have reported the relationship between OPLL and various other diseases and traits, especially type 2 diabetes (T2D) (Akune et al., 2001; Kobashi et al., 2004), high body mass index (BMI) (Hou et al., 2017; Kobashi et al., 2004), low inorganic phosphate, X-linked hypophosphatemic rickets (Chesher et al., 2018), and increased C-reactive protein (Kawaguchi et al., 2017). Of these, T2D has been the focus of attention for a long time (Akune et al., 2001; Kobashi et al., 2004). Furthermore, genetic factors have long been considered in OPLL development (Matsunaga et al., 1999; Sakou et al., 1991; Terayama, 1989). To understand the genetic factors associated with OPLL, we previously conducted a genome-wide association study (GWAS) and found six significant loci (Nakajima et al., 2014). In subsequent *in silico* and *in vitro* functional studies, *RSPO2* has been identified as a susceptibility gene for OPLL, and the role of Wnt signaling in the pathogenesis of OPLL is disclosed (Nakajima et al., 2016). However, the pathogenesis of this condition remains unknown.

In this study, to clarify the etiology and pathogenesis of OPLL, we conducted a meta-analysis of GWASs and various post-GWAS analyses. We identified 14 significant loci, including eight previously unreported susceptibility loci. Using a gene-based analysis (de Leeuw et al., 2015) and summary data-based Mendelian randomization (SMR) (Zhu et al., 2016), we identified three significant risk genes outside the GWAS significant region. Using a genetic correlation analysis and a subsequent Mendelian randomization (MR) study, we identified a causal effect of high BMI on OPLL. A polygenic risk score (PRS) of BMI demonstrated the heterogeneity of the impact of obesity on OPLL subtypes.

## Results

### Novel susceptibility loci in OPLL

We conducted three GWASs (set 1-3) in the Japanese population (Supplementary Table 1). After quality control of single nucleotide polymorphism (SNP) genotyping data, we performed imputation and association analyses independently for each GWAS. Subsequently, we performed a fixed-effects meta-analysis combining the three GWASs (ALL-OPLL: a total of 2,010 cases and 20,006 controls; Supplementary Fig. 1) and identified 12 genome-wide significant loci (*P* < 5.0×10^−8^) (Fig. 1). The genomic inflation factor (λGC) was 1.11 and showed slight inflation in GWAS; however, the intercept in linkage disequilibrium (LD) score regression (Bulik-Sullivan et al., 2015) was 1.03, indicating that inflation of the statistics was mostly from polygenicity and minimal biases of the association results (Supplementary Fig. 2).

**Fig. 1.**
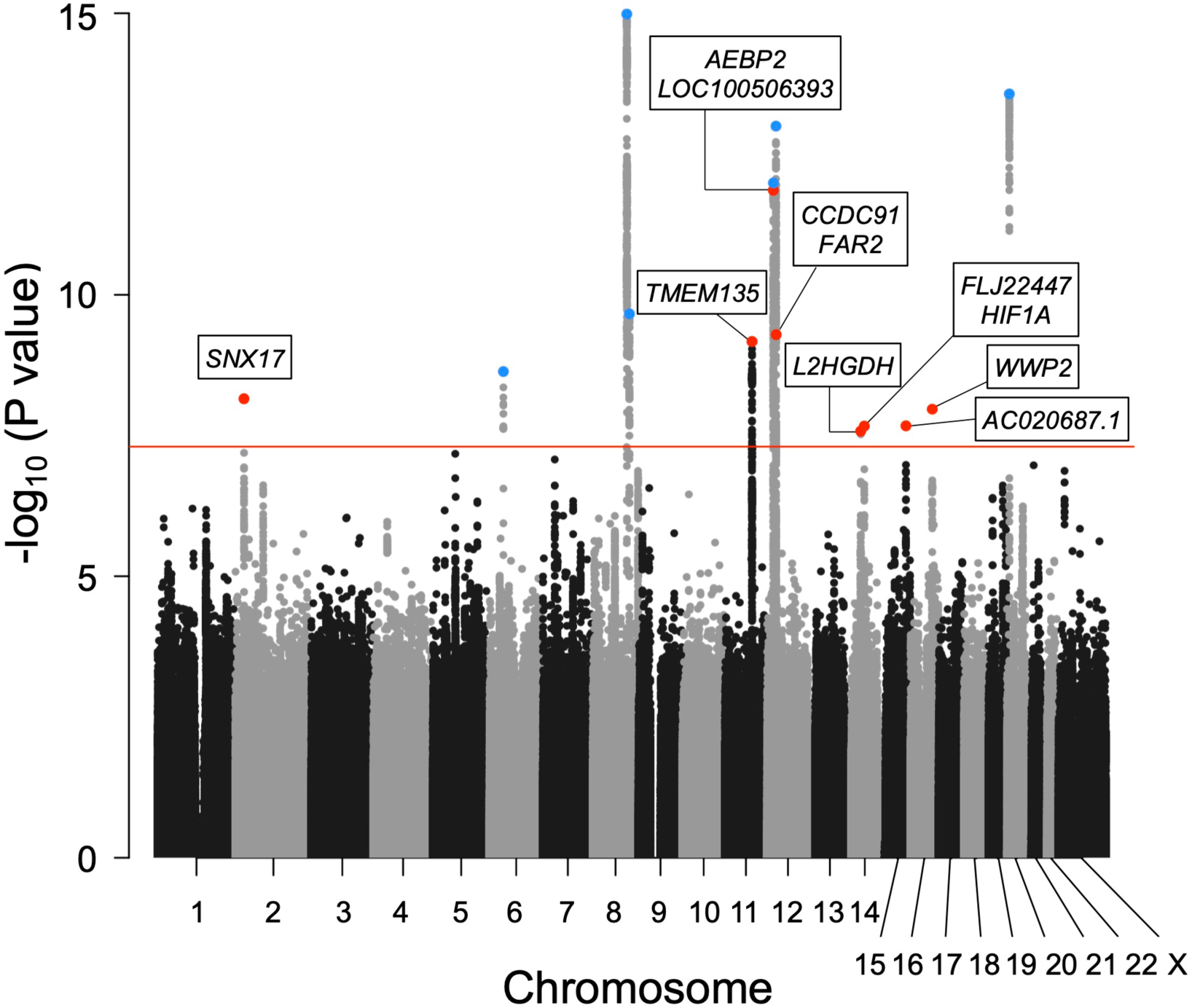
Meta-analysis of GWAS identified 14 significant loci in OPLL Manhattan plot showing the -log10 P-value for each SNP in the meta-analysis. The values were plotted against the respective chromosomal positions. The horizontal red line represents the genome- wide significance threshold (*P* = 5.0 × 10^-8^). Red and blue points represent the lead SNPs in the new and known loci, respectively.

Next, we conducted a stepwise conditional analysis to detect multiple independent signals. We detected two additional independent signals that showed genome-wide significance after conditioning (Supplementary Table 2): rs35281060 (12p12.3, *P* = 1.44×10^−12^) and rs1038666 (12p11.22, *P* = 5.00×10^−10^) (Supplementary Fig. 3f-i). We also detected one additional signal (rs61915977, 12p11.22 *P* = 1.39×10^−6^) that reached locus-wide significance (*P* < 5.0 × 10^-6^) (Supplementary Table 2). Thus, the meta-analysis and conditional analysis identified 14 genome-wide significant OPLL loci, including eight novel loci. Significant associations of the six previously reported loci (Nakajima et al., 2014) were observed in the present study (Table 1, Fig. 1, Supplementary Fig.3). The estimated proportion of the phenotypic variance explained by all the variants used in the study was 53.1% (95% confidence interval (CI): 40.6 - 65.6%), indicating that OPLL has a high heritability. The lead variants of the 14 loci explained 6.5% of the phenotypic variance. Together with the results of the LD score regression, OPLL is a highly polygenic disease.

**Table 1.**
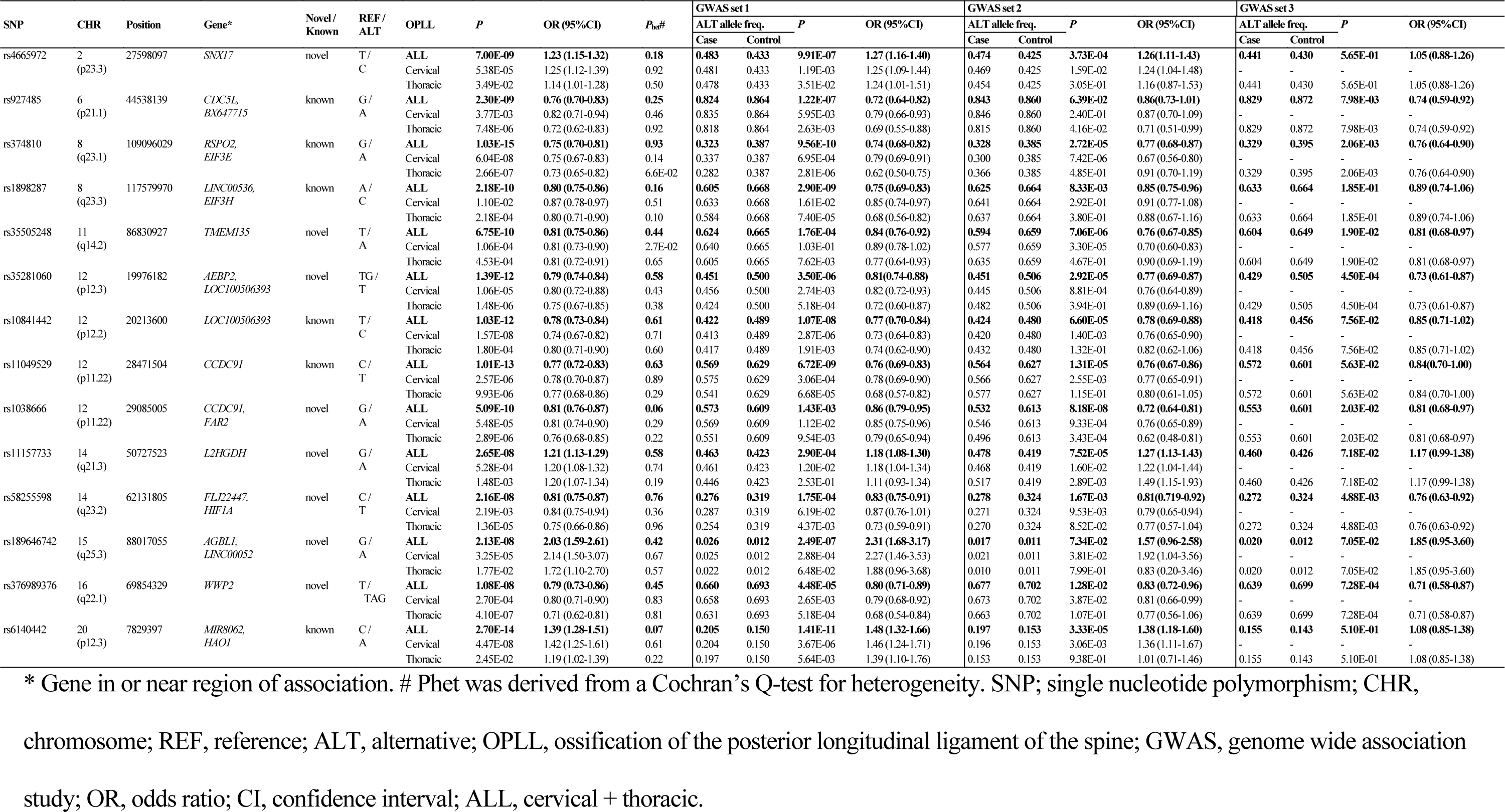
Genome-wide significant loci in ossification of the posterior longitudinal ligament of the spine

Adjacent to lead variants in the novel loci, we found several candidate causal genes (Fig. 1), reported to be related to osteogenesis and could be connected to OPLL development. *TMEM135* (transmembrane protein 135), a gene in the newly identified significant locus (11q14.2), is a multi-transmembrane protein with seven transmembrane helices of high confidence. It is more strongly expressed in multipotent adipose tissue-derived stem cells committed to osteoblastic cells than the adipogenic lineage (Scheideler et al., 2008). *WWP2* (WW domain-containing E3 ubiquitin-protein ligase 2), the nearest gene to rs376989376 (the lead SNP in 16q22.1), was recently reported to serve as a positive regulator of osteogenesis by augmenting the transactivation of *RUNX2*, a master regulator of osteoblast differentiation as well as for chondrocyte maturation during skeletal development (Zhu et al., 2017).

All lead SNPs and SNPs in high LD (r^2^ > 0.8) with them in previously unreported significant loci were in intron or intergenic regions, and none of them were exonic variants (Supplementary Table 3). To prioritize putative causal variants, we conducted a Bayesian statistical fine-mapping analysis for significant loci using FINEMAP (Benner et al., 2016). The lead SNPs had the highest posterior probability (PP) in any significant region except 20p12.3, and two of them were higher than 0.5: rs4665972 (2p23.3, PP = 0.541) and rs1038666 (12p11.22, PP = 0.545) (Supplementary Table 4).

### Enrichment in genes involved in bone metabolism

We conducted a gene set enrichment analysis implemented in FUMA (Watanabe et al., 2017). We found significant enrichment in the set related to bone mineral density (BMD): BMD of the heel (*P=* 8.60 ×10^−8^), pediatric lower limb (*P*= 9.24 ×10^−5^), and pediatric total body less head (*P*= 2.68 ×10^−4^) (Supplementary Table 5), compatible with the critical roles of bone metabolism in OPLL.

### Identification of novel candidate genes missed by the GWAS meta-analysis

To identify other potential causal genes, we conducted a gene-based association analysis (de Leeuw et al., 2015; Watanabe et al., 2017). We found five additional genes significantly associated with OPLL: *SFRP4, DLX6, EIF3E*, *EMC2*, and *TMEM135* (Fig. 2, Supplementary Table 6). Of these genes, *SFRP4* (secreted frizzled-related protein 4) and *DLX6* (distal-less homeobox 6), located on chromosome 7 (p14.1 and q21.3, respectively), seemed to be plausible causal genes of OPLL. *SFRP4* is the gene whose loss-of-function mutations cause Pyle’s disease (OMIM 265900), a rare autosomal recessive skeletal dysplasia characterized by wide metaphyses with increased trabecular bone, significant cortical thinning, fractures, and thin calvarium (K. Chen et al., 2019; Kiper et al., 2016). It has been reported that *Dlx5* and *Dlx6* are functionally redundant regulators of chondrocyte hypertrophy (Hsu et al., 2006), and the absence of *DLX5* and *DLX6* causes split hand/split foot malformation type I (Rattanasopha et al., 2014).

**Fig. 2.**
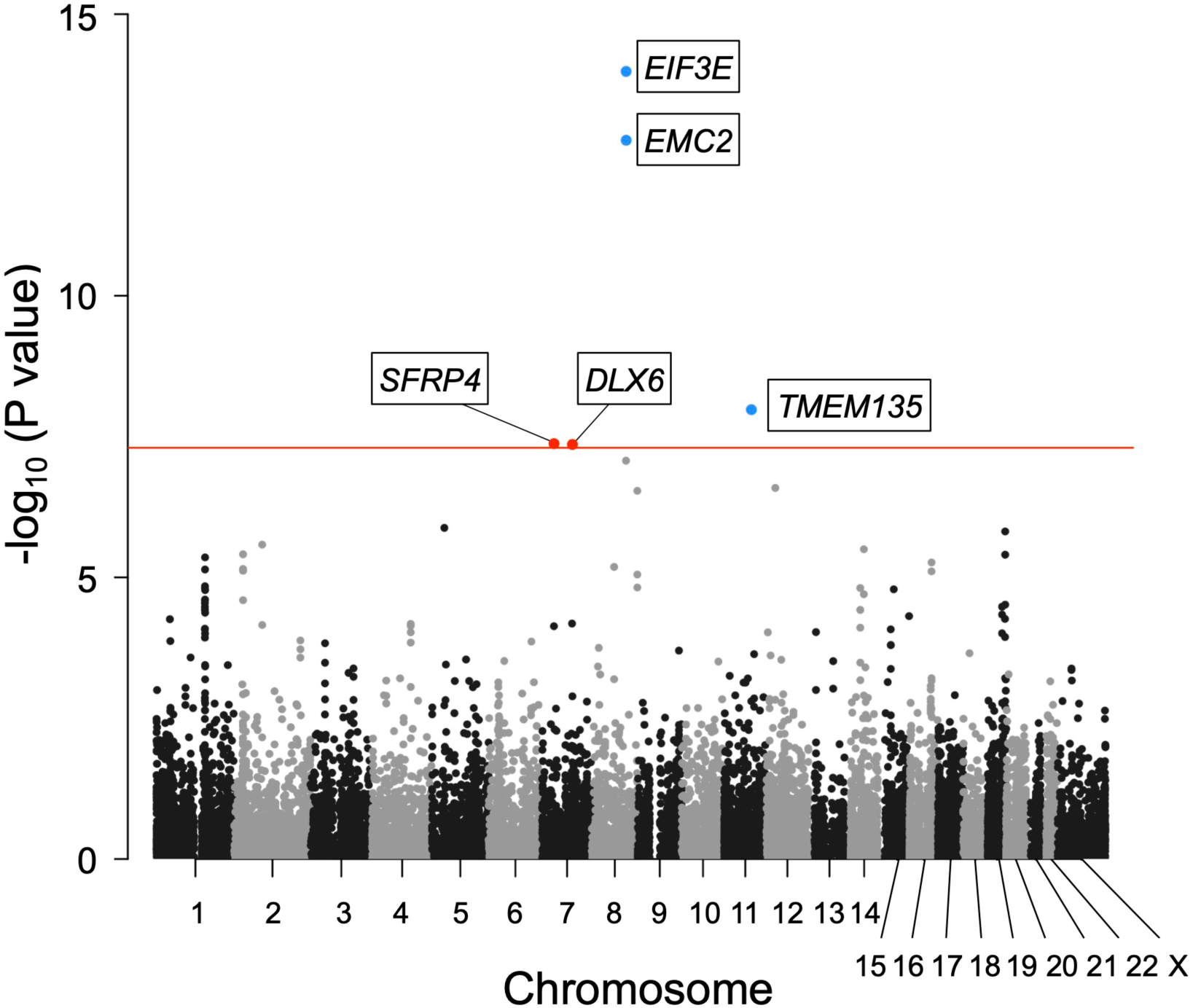
Gene-based association analysis identified five significantly associated genes in OPLL. Manhattan plot showing the -log10 P-value for each gene in the analysis. The values were plotted against the respective chromosomal positions. The horizontal red lines represent significance threshold (*P* = 5.0 × 10^-8^). Red and blue points represent the significant risk genes outside and inside the GWAS-significant region, respectively.

The lack of exonic variants suggests that altering gene expression levels is a key function of OPLL-associated variants. By searching eQTL data in all available tissues in GTEx (GTEx Consortium, 2015), we found 26 transcripts with cis-eQTL variants associated with OPLL signals; of these, 20 transcripts were in the novel loci (Supplementary Table 7). Furthermore, SMR (Zhu et al., 2016) revealed a total of 10 gene-tissue pairs (three unique genes, namely, *RSPO2*, *PLEC*, and *RP11-967K21.1*) that surpassed the genome-wide significance level (*P*SMR < 8.4×10^-6^) without heterogeneity (*P*HEIDI < 0.05) (Supplementary Table 8). *RSPO2* is located in the most significant locus in GWAS meta-analysis, and its functions related to OPLL were elucidated in a past study (Nakajima et al., 2016). *PLEC* is expressed in various tissues, including muscles and fibroblasts (GTEx Consortium, 2015), and *PLEC* deficiency causes epidermolysis bullosa simplex with muscular dystrophy (OMIM 226670)(Smith et al., 1996), in which osteoporosis frequently develops (J. S.-C. Chen et al., 2019). Since increased expression of *PLEC* was estimated to have a causal effect on OPLL (Supplementary Fig. 4, Supplementary Table 8), these results suggest that *PLEC*is a likely causal gene of OPLL.

### Cell group and cell types related to OPLL

We conducted cell type group enrichment analysis of the polygenic signals to investigate cell groups related to OPLL. We observed significant enrichment in the active enhancers of the connective/bone cell group (Supplementary Table 9). We then analyzed each cell type belonging to the group and found significant enrichment of H3K27ac in chondrogenic differentiation cells (Supplementary Table 10). These results are concordant with previous findings that in OPLL, chondrocyte differentiation in the endochondral ossification process occurs (Sugita et al., 2013).

### Subtype analyses of OPLL

Subtype-stratified GWAS meta-analyses were also conducted: cervical (C)-OPLL (820 cases and 14,576 controls) and thoracic (T)-OPLL (651 cases and 20,007 controls). Subsequently, we identified three significant loci for C-OPLL (Supplementary Fig. 5, 6, 7, Supplementary Table 11) and eight significant loci for T-OPLL (Supplementary Fig. 5, 6, 8, Supplementary Table 11). Of these loci, one in the C-OPLL analysis and eight in the T-OPLL analysis were not identified in the analysis of ALL-OPLL and other OPLL subtypes. We identified one plausible locus among the T- OPLL susceptibility signals. rs74707424, a leading SNP in the significant locus (19p12), is located in the 3’-untranslated region of the *ZBTB40* gene. In a recent study using primary osteoblasts of mouse calvaria, Doolittle *et al*. reported that *Zbtb40* functions as a regulator of osteoblast activity and bone mass, and knockdown of *Zbtb40,* but not *Wnt4*, in osteoblasts drastically reduced mineralization (Doolittle et al., 2020). We did not find any significant genes in the gene-based analysis (data not shown).

### Causality of high BMI on OPLL

Epidemiological studies have suggested a relationship between OPLL and various other diseases and traits (Akune et al., 2001; Endo et al., 2020; Kawaguchi et al., 2017; Kobashi et al., 2004), particularly with T2D (Akune et al., 2001; Kobashi et al., 2004). We investigated their relationship with OPLL using the GWAS data. We first calculated the genetic correlation between OPLL and 99 complex traits (42 diseases and 57 quantitative traits, mean N of 130,582) (Akiyama et al., 2017; Ishigaki et al., 2020; Kanai et al., 2018). We found a positive genetic correlation between OPLL and BMI and T2D. The genetic correlation estimate (rg) was higher in the BMI group than in the T2D group (Fig. 3). In addition, we identified new negative correlations between cerebral aneurysms and osteoporosis (Fig. 3, Supplementary Table 12). This negative correlation between OPLL and osteoporosis is concordant with the fact that OPLL patients have an increased tendency of ossification throughout the body (Mori et al., 2016; Nishimura et al., 2018; Yoshii et al., 2019).

**Fig. 3.**
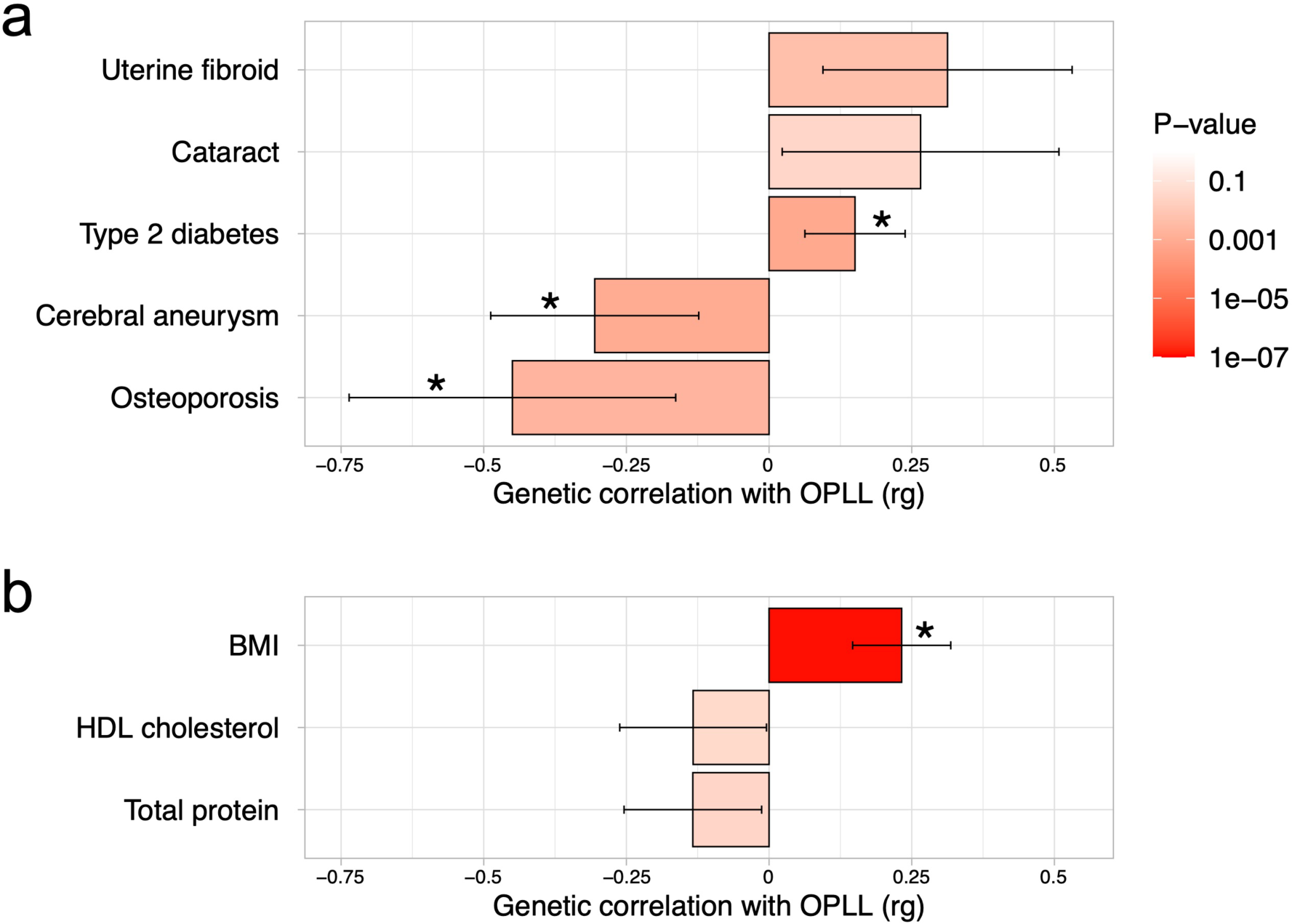
Genetic correlation between OPLL and other complex traits (a) Disease. (b) Quantitative trait. Significant positive correlations with BMI and type 2 diabetes, and negative correlations with cerebral aneurysm and osteoporosis were observed. Error bars indicate 95% confidence intervals. Red color gradations represent the level of P-value. Noted by asterisk is the significant correlation (FDR <0.05).

Next, we conducted a two-sample MR using summary data from GWASs (Akiyama et al., 2017; Bakker et al., 2020; Kemp et al., 2017; Spracklen et al., 2020) to assess the causal effects of these traits on OPLL (Evans and Davey Smith, 2015; Lawlor et al., 2008) (Supplementary Fig. 9, Supplementary Table 13). The significant causal effect of genetically increased BMI on ALL-OPLL was estimated using the inverse variance weighted method and weighted median method (Fig. 4, Supplementary Fig. 10, Supplementary Table 14). The average pleiotropic effect of the MR-Egger regression intercept was close to zero (MR-Egger intercept = 0.005, P-value= 0.580), indicating no evidence of the influence of directional pleiotropy (Supplementary Fig. 10, Supplementary Table 15). We also assessed potential bias in the MR with a leave-one-out analysis and funnel plots; however, we did not identify any obvious bias (Supplementary Fig. 11). In contrast, we could not find any causal effects of T2D on ALL-OPLL with any MR methods (Fig. 4, Supplementary Fig. 12, Supplementary Table 14). As for osteoporosis, characterized by a loss of BMD, we found a weak but significant causal effect of genetically increased BMD on ALL-OPLL using multiple MR methods (Fig. 4, Supplementary Fig. 13, Supplementary Table 14), and the involvement of factors that stimulate bone formation in OPLL was suggested. Regarding cerebral aneurysms, the direction of the beta estimates differed according to the MR methods because of the small number of SNPs used as the instrumental variables (Fig. 4, Supplementary Fig. 14, Supplementary Table 14).

**Fig. 4.**
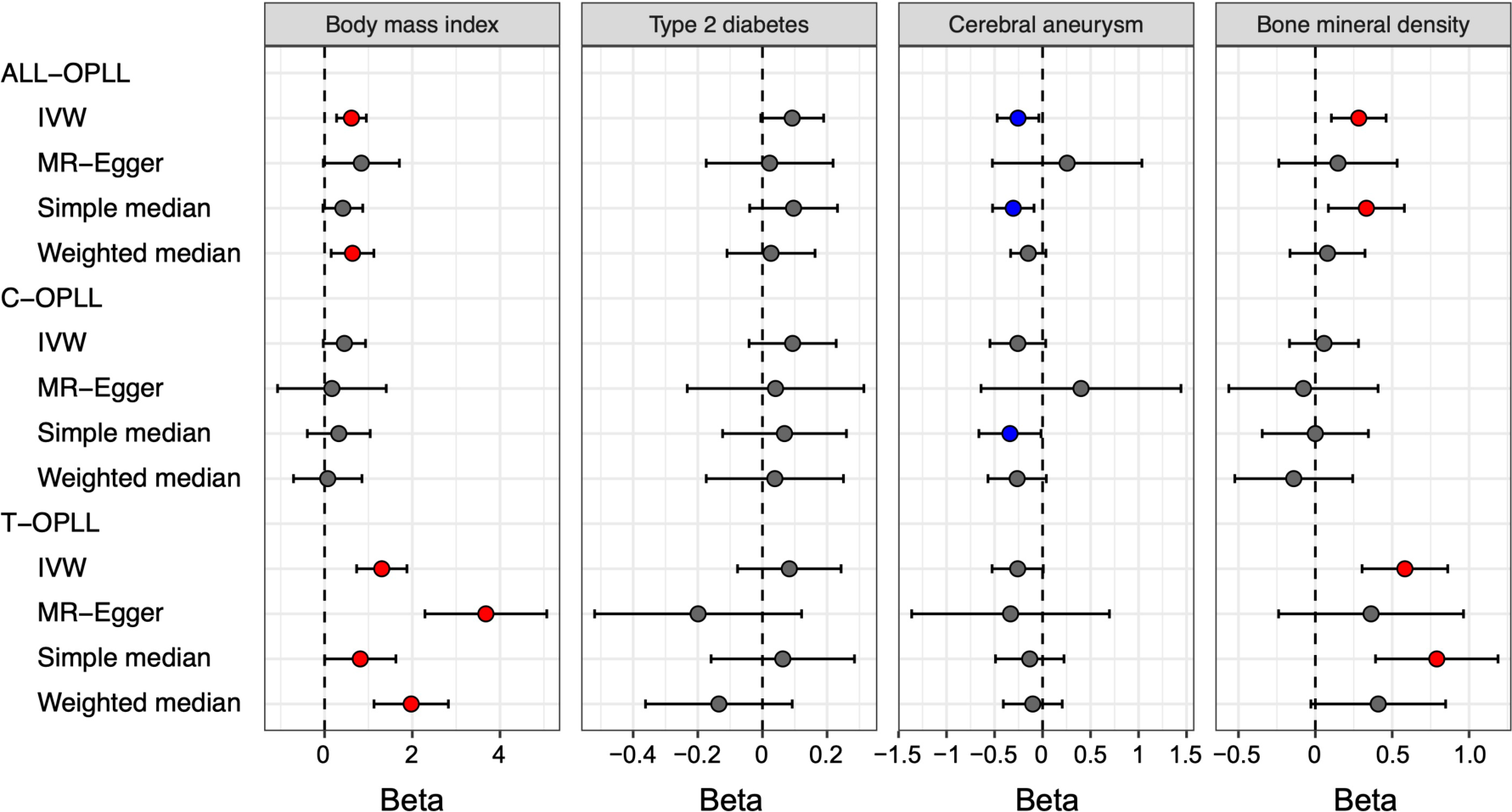
Causal effect of body mass index, type 2 diabetes, cerebral aneurysm, and bone mineral density on OPLL Causal effects were estimated using two-sample MR methods. Error bars indicate 95% confidence intervals. Significant (*P* < 0.05) results are shown as red and blue dots for positive and negative causal effects, respectively. IVW, inverse variance weighted.

We performed a reverse-direction MR to evaluate the causality of OPLL on BMI, T2D, cerebral aneurysm, and BMD but did not find any significant causal effects on the four traits (Supplementary Fig. 15, Supplementary Tables 16 and 17).

**Fig. 5.**
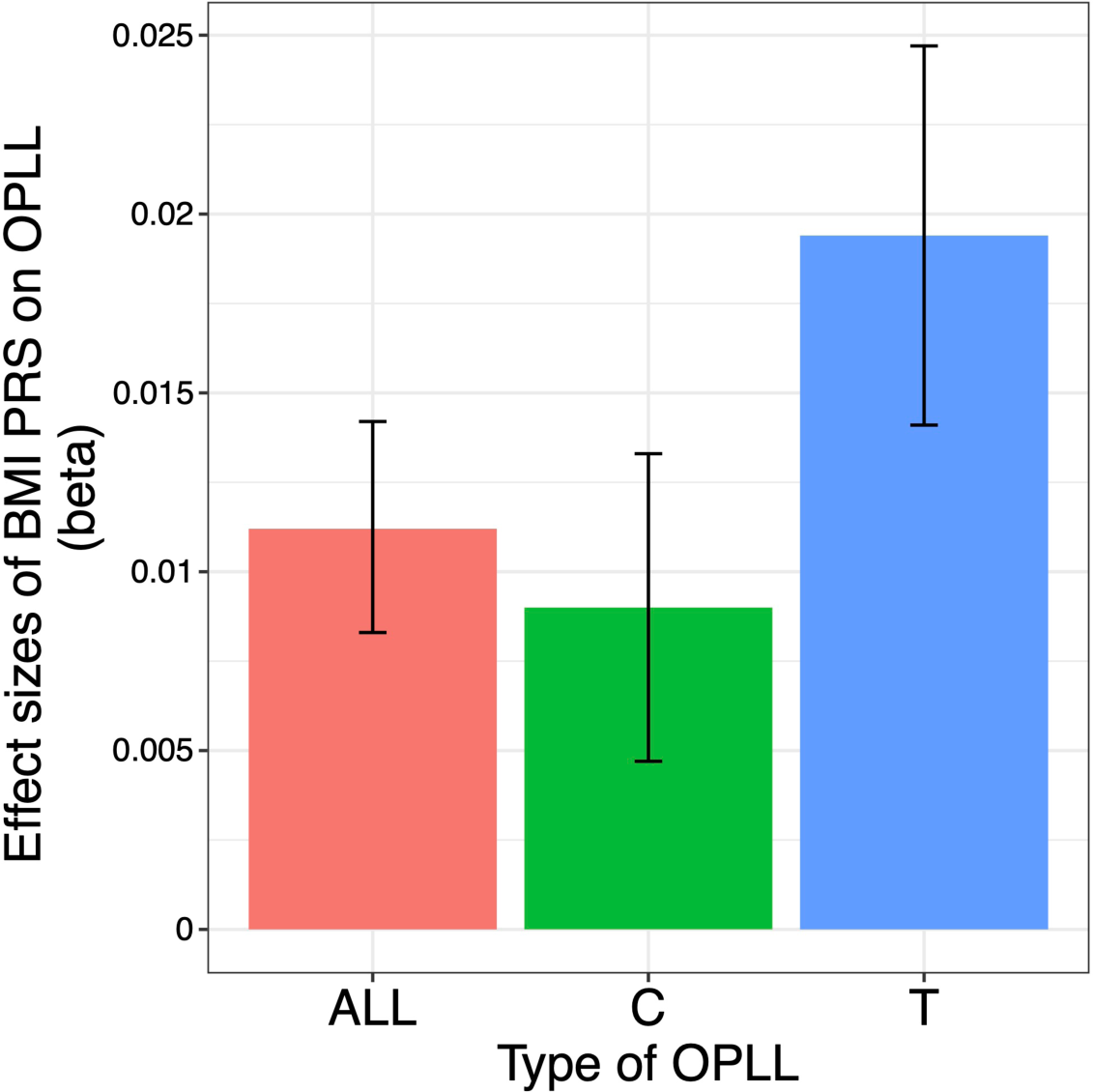
BMI polygenic risk score predicts OPLL Vertical columns represent effect sizes of BMI PRS on three types of OPLL: cervical (C-) OPLL, thoracic (T-) OPLL, and All-OPLL (C-OPLL+T-OPLL). The BMI PRS could predict OPLL, especially T-OPLL. Error bars represent the 95% confidence intervals of the effects.

### The large causal effect of high BMI and high BMD on T-OPLL

We estimated the causal effect of traits that had a significant genetic correlation with OPLL subtypes. We found contrasting results between C- and T-OPLL. A significant causal effect of genetically increased BMI on T-OPLL, but not on C-OPLL, was indicated by all four MR methods. All beta estimates on T-OPLL were greater than those in the analysis for ALL-OPLL (Fig. 4, Supplementary Fig. 10, 16, Supplementary Table 14), suggesting that T-OPLL drove the causal effect of BMI on OPLL. The MR-Egger regression intercept was significantly negative, suggesting the existence of directional pleiotropy (Supplementary Fig. 16, Supplementary Table 15); however, the results of other sensitivity analyses for robust causal inference (simple and weighted median methods) suggested its causality on T-OPLL (Fig. 4, Supplementary Fig. 16 and 17, Supplementary Table 14). As for BMD, a larger causal effect of genetically increased BMD on T-OPLL compared to ALL-OPLL was also estimated (Fig. 4, Supplementary Fig. 13, Supplementary Table 14). We did not find any significant effects in T2D or cerebral aneurysms except for the results using simple median method for cerebral aneurysms.

### The polygenic causal effect of high BMI on OPLL

In addition to the causal effects of significant variants with BMI on OPLL, we evaluated the shared polygenic architecture between BMI and OPLL. Moderate correlations were found between the effect sizes of the SNPs of ALL- and T-OPLL with BMI, especially in sets of SNPs with low P-values (*P* < 0.0005), when calculating the correlation based on P-value in the BMI GWAS. We did not find such correlations when calculating the correlation based on the P-value in the OPLL GWAS (Supplementary Fig. 18). However, this difference was not observed in the sets of SNPs of any P-value groups in the C-OPLL analysis. These results support the theory that high BMI is one of the causal factors of OPLL, and its causal effect on OPLL is driven by that on T-OPLL.

### Heterogeneity of impact of obesity inside OPLL subtypes

Finally, we generated a PRS of BMI and compared its effect on OPLL for ALL-OPLL, C-OPLL, and T-OPLL (Methods, Supplementary Fig. 19, 20). We found that BMI-PRS could predict OPLL, especially T-OPLL (Fig. 5). Comparing C- and T-OPLL, we found that the effect of BMI PRS was significantly greater in T-OPLL (*P* = 0.016), even in a limited number of cases.

## Discussion

This study conducted a GWAS meta-analysis using 2,010 OPLL cases and 20,006 controls and identified 14 significant loci, including eight novel loci. The association signals of OPLL converged in every aspect to activate bone formation and suppress bone resorption and showed significant enrichment in the gene set related to bone metabolism. In addition, we identified additional genes associated with OPLL using gene-based analysis and an SMR: *SFRP4*, *DLX6*, and *PLEC*. Chen *et al*. reported osteoblast- and osteoclast-expressed *Sfrp4* regulates osteoclast differentiation and bone resorption activity via noncanonical Wnt/Ror2/Jnk signaling in osteoclasts (K. Chen et al., 2019) *Dlx6* is expressed at high levels in osteoblasts and has a potential to stimulate osteoblastic differentiation (Li et al., 2008). The *PLEC* mutation causes epidermolysis bullosa, in which osteoporosis is one of the main comorbidities (J. S.-C. Chen et al., 2019). Thus, our GWAS observations are compatible with the theory that OPLL is closely related to bone metabolism and develops through the process of endochondral ossification (Sato et al., 2007; Sugita et al., 2013).

We identified that OPLL was genetically correlated with other common diseases, positive for BMI and T2D and negative for osteoporosis and cerebral aneurysms. High BMI (obesity) has been implicated in OPLL, and clinical studies have reported that the BMI of OPLL patients is significantly higher than that of non-patients (Endo et al., 2020; Kobashi et al., 2004). It is speculated that obesity promotes ectopic bone formation in OPLL. Yamamoto *et al*. reported a high incidence of OPLL in the Zucker fatty rat, a model mouse of obesity with a loss-of-function mutation in the leptin receptor gene (Yamamoto et al., 2004). Lv *et al*. reported that a high-fat diet (HFD)-induced obesity promotes bone formation. BMD and trabecular thickness at the femur were significantly greater in HFD-induced obese mice than in normal control mice. The mRNA level of *Runx2* in bone marrow-derived mesenchymal stem cells from HFD mice was significantly higher. In contrast, that of *PPARγ*, which suppresses osteoblast differentiation and promotes osteoclast differentiation, was significantly lower than the control mice (Lv et al., 2010). In addition, MR analysis of the relationship between BMI and BMD reported that increased BMI was positively causally related to heel BMD (Ma et al., 2021; Song et al., 2020) and lumbar BMD (Song et al., 2020), supporting the theory that obesity is related to OPLL formation. Our MR studies have demonstrated that a high BMI has a causal effect on OPLL. However, we could not prove the SNP-obesity interaction (Supplementary Note 2). Further mechanistic studies on obesity and OPLL are necessary, which would lead to novel non-surgical treatment and prevention of OPLL.

We did not identify any causal effects of T2D on OPLL by our MR analysis, despite the positive genetic correlation between OPLL and T2D. Many OPLL studies have focused on the relationship with T2D (Akune et al., 2001; Kobashi et al., 2004). Insulin acts on endogenous tyrosine kinase receptors and receptors for insulin-like growth factor-I, a potent anabolic factor in bone formation (Giustina et al., 2008; Locatelli and Bianchi, 2014). Therefore, it has been postulated that increased insulin production due to impaired insulin action may stimulate osteogenic cells in ligaments and cause OPLL (Akune et al., 2001). However, our results indicate that the impact of T2D on the development of OPLL is not significant. Therefore, most of the reasons for the high prevalence of T2D in OPLL patients reported so far can be attributed to the high prevalence of obesity in OPLL patients.

Our MR analysis also revealed the causality of high BMD in OPLL. This is consistent with the clinical observation that OPLL patients have an increased tendency of ossification throughout the body and often have ectopic ossification of other spinal ligaments, such as the anterior longitudinal ligament (Nishimura et al., 2018), interspinous ligament (Mori et al., 2016), and nuchal ligament (Yoshii et al., 2019), as well as extraspinal ossification lesions in the shoulder, hip, knee, and ankle joint (Hukuda et al., 1983). However, the relationship between cerebral aneurysms and OPLL has not been reported. Therefore, for evaluating the causality of cerebral aneurysms on OPLL, an MR analysis with more cerebral aneurysm-related SNPs as the instrumental variables is desirable. Such a study will be conducted in the future.

We demonstrated the differences in genetic characteristics of OPLL. We identified three significant loci for C-OPLL and eight significant loci for T-OPLL in the stratification analyses of the GWAS by OPLL subtypes. There was a considerable difference in the allele frequencies of lead variants in these loci between subtypes (Supplementary Table 11). However, most of the alternative allele frequencies of these variants were small, and future confirmation by a study with larger sample size is desirable. Furthermore, our MR and BMI PRS analyses showed that the effect of BMI on T- OPLL was much larger than that of C- and ALL-OPLL. The common disease study often shows clinically defined diseases based on common signs and symptoms are actually heterogeneous in the cause, such as hypertension and diabetes mellitus. Therefore, research focusing on disease subtypes is useful in characterizing the disease in detail and elucidating its specific causes, leading to more personalized treatment.

## Materials and methods

### Subjects

All the subjects analyzed in this study were Japanese. The GWAS data of this study consisted of three sets: GWAS set-1, -2, and -3. The case samples of set-1 and -2 were used as discovery and replication samples, respectively, in the previous GWAS (Nakajima et al., 2014). In these data sets, the cases had OPLL of more than or equal to two vertebrae. For the case of set-3, we recruited patients with OPLL in more than or equal to 5 vertebras or OPLL thicker than 5 mm in the thoracic spine in 2018-2019. When assessing the presence or absence of OPLL, expert spine surgeons in each institution examined patients’ plain radiography or computed tomography (CT) in detail (Supplementary Fig. 1).

Regarding control data, we used genotyping data from BioBank Japan (BBJ) (Hirata et al., 2017; Nagai et al., 2017) in set-1and -2, and those from the Medical Genome Center (MGC) Biobank database of the National Center for Geriatrics and Gerontology (NCGG) in set-3. Details of the characteristics of the subjects are shown in Supplementary Table 1.

This study followed the Strengthening the Reporting of Genetic Association Studies (STREGA) reporting guideline (Little et al., 2009).

### Study approval

All participating individuals provided written informed consent to participate in this study following approval by the ethics committees of the participating institutions (Supplementary Data).

### Genotyping and quality control

Genomic DNA was extracted from peripheral venous blood samples using a standard method. We genotyped case and control samples using the Illumina OmniExpressExome BeadChip, a combination of Illumina OmniExpress BeadChip and Illumina HumanExome BeadChip, or Illumina Asian Screening Array (Supplementary Table 1).

For quality control of genotyped SNPs, we excluded those with (i) SNP call rate < 99%, (ii) minor allele frequency (MAF) < 0.01, and (iii) Hardy-Weinberg equilibrium (HWE) P*-*value < 1.00 × 10^-6^. We constructed a reference panel to obtain imputed genotypes with high accuracy using the 1000 Genomes Project Phase 3 [1KGP 3 (May 2013, n = 2,504)] and 3,256 in-house Japanese whole-genome sequence data obtained from BBJ (JEWEL 3K) in the same way as previously reported (Akiyama et al., 2019). SNPs with allele frequency differences greater than 0.06 between the genotyped control data and the 1KGP3 East Asian and JEWEL 3K data in reference panel were excluded.

For sample quality control, we excluded samples whose sex differed between genotype and clinical data. We evaluated cryptic relatedness by calculating estimates of pairwise IBD (PI_HAT) and removed samples that showed second-degree relatedness or closer (PI_HAT > 0.25). Population stratification was estimated using principal component analysis (PCA) with four populations from HapMap data as the reference: European (CEU), African (YRI), Japanese (JPT), and Han Chinese (CHB) with SmartPCA (Patterson et al., 2006). We generated a scatterplot using the top two associated principal components (eigenvectors) and selected samples within the East Asian (JPT/CHB) cluster. We excluded samples with a genotyping call rate of < 98% (Supplementary Fig. 1).

### Phasing and genotype imputation

We performed pre-phasing with EAGLE (v2.4.1) (Loh et al., 2016) and SNP imputation with minimac4 (v1.0.0) (Das et al., 2016) using the reference panel mentioned above. After imputation, we used SNPs with an imputation quality of Rsq > 0.3 and MAF > 0.005 for the subsequent association study.

### GWAS and meta-analysis

We performed an association analysis of autosomes of GWAS set-1, -2, and -3 independently. We performed a logistic regression analysis using PLINK2.0 (Purcell et al., 2007) with adjustment for 10 principal components, assuming an additive model and evaluated the association of each imputed SNP. We then meta-analyzed the three GWAS sets with an inverse-variance method under a fixed effect model using METAL software (Willer et al., 2010). Regarding X chromosomes, we performed a logistic regression analysis in males and females separately for each GWAS set using PLINK2.0, with adjustment for 10 principal components assuming an additive model. We then integrated the results of males and females in each GWAS using an inverse-variance method under a fixed-effect model (Supplementary Fig. 1).

We calculated LD with the lead variants using whole-genome sequence data of 1KGP3 East Asian and JEWEL 3K by PLINK2.0 and produced regional association plots using Locuszoom (Pruim et al., 2010) (http://locuszoom.org). We estimated confounding biases derived from population stratification and cryptic relatedness using LD score regression using LD scores for the East Asian population (Bulik-Sullivan et al., 2015).

To identify candidate causal variants in the eight novel loci, we annotated the SNPs that exceeded the threshold of significance (*P* < 5.0×10^−8^) and were in high LD (*r*^2^ > 0.8) with lead variants newly identified in the GWAS meta-analysis. We explored the biological role of these variants using SNP annotation tools, including HaploReg (Ward and Kellis, 2012) (https://pubs.broadinstitute.org/mammals/haploreg/haploreg.php), RegulomeDB (Boyle et al., 2012) (http://regulomedb.org/), and ANNOVAR (Wang et al., 2010) (http://annovar.openbioinformatics.org/en/latest/).

### Subtype-stratified GWAS and meta-analysis

We performed subtype-stratified GWASs and meta-analyses for C-OPLL and T-OPLL in the same way as the analysis with all samples from set to 1-3 (ALL-OPLL). Cases in GWAS set-3 were all T-OPLL samples; therefore, we carefully re-examined patients’ plain radiography or CT in set-1 and -2, where we defined C-OPLL case samples as those with OPLL limited to the cervical spine, and defined T-OPLL as OPLL affecting more than two vertebrae in the thoracic spine (Supplementary Fig. 1). The detailed sample numbers are listed in Supplementary Table 1.

### Conditional association analysis

We defined an associated locus of a lead variant as 1 Mb of its surrounding sequences in both directions. We extended the region to nearby significant variants and their 1 Mb surrounding sequences as far as a significant variant was contained in the defined region. We performed a stepwise conditional meta-analysis to determine the independent association signals in the associated loci. We conducted conditional analyses of GWAS set 1-3 separately and integrated the results using a fixed-effects model with the inverse variance weighted method. We repeated this process until the top associated variants fell below the locus-wide significance level (*P* < 5.0 × 10^-6^) in each stepwise procedure.

### Estimation of phenotypic variance

We estimated the heritability of OPLL using LDSC software (Bulik-Sullivan et al., 2015). The variance explained by the variants was calculated based on a liability threshold model by assuming the prevalence of OPLL to be 3.0% (Matsunaga and Sakou, 2012). The model assumed that subjects had a continuous risk score and that subjects whose scores exceeded a certain threshold developed OPLL.

### Bayesian statistical fine-mapping analysis

To prioritize causal variants in OPLL susceptibility loci, we conducted a fine-mapping analysis using FINEMAP v1.3 software (Benner et al., 2016), using z-scores of GWAS meta-analysis for ALL-OPLL and LD matrices calculated by 1KGP3 EAS and JEWEL 3K data. We assumed one causal signal in the ±1 Mb region from both ends of significant variants at each significant locus, but for 12p11 and 12p12, in which we identified significant secondary signals by a conditional analysis, we defined the range of the region referring to regional association plots (Supplementary Fig. 3). We calculated a PP in which each genetic variant was the true causal variant. We ranked the candidate causal variants in descending order of their PPs and created a 95% credible set of causal variants by adding the PPs of the ordered variants until their cumulative PP reached 0.95.

### Gene set enrichment analysis

We conducted a gene set enrichment analysis using FUMA (Watanabe et al., 2017). We selected the genes based on the following criteria and used them as input data: genes (i) located within 1 Mb and (ii) the five closest to the leading SNPs of each genome-wide significant locus.

### Gene-based association analysis

To examine the combined effect of SNPs, we conducted gene-based association analysis using MAGMA (de Leeuw et al., 2015) implemented in FUMA (Watanabe et al., 2017). We used the default settings and LD information from East Asian ancestry subjects from 1KGP3 as a reference. We set the gene window 2 kb upstream and 1 kb downstream from the genes to include regulatory elements and analyzed 19,737 genes. We set the P-value threshold for the test to 5.0 × 10^-8^ (not a gene-wide threshold).

### eQTL analysis

We obtained transcript data from the Genotype-Tissue Expression (GTEx) v8 (GTEx Consortium, 2015). We examined eQTL data in all available tissues in GTEx to determine the association between gene expression and the leading SNPs within the genome-wide significant locus. We set the significance threshold for eQTL as a false discovery rate (FDR) < 0.05.

### SMR

We used SMR software (Zhu et al., 2016). We used OPLL summary statistics data and eQTL data obtained from GTEx v7 (GTEx Consortium, 2015). We evaluated heterogeneity in dependent instruments (HEIDI) using multiple SNPs in a *cis*-eQTL region to distinguish pleiotropy from linkage. We set the threshold for the HEIDI test to 0.05 and the threshold for SMR to 8.4 × 10^-6^ as previously reported (Zhu et al., 2016).

### Cell-type-specific enrichment analysis

We performed stratified LD score regression using 220 cell-type-specific annotations of four histone marks (H3K4me1, H3K4me3, H3K9ac, and H3K27ac) (Roadmap Epigenomics Consortium et al., 2015). We divided the 220 cell-type-specific annotations into ten cell type groups (10 in adrenal/pancreas, 34 in central nervous system, 15 in cardiovascular, 6 in connective/bone, 44 in gastrointestinal, 67 in immune/hematopoietic, 5 in kidney, 6 in liver, 10 in skeletal muscle, and 23 in other). We assessed heritability enrichment in histone marks of 220 individual cell types and ten cell type groups, as described by Finucane et al. (Finucane et al., 2015). The regression analysis excluded variants within the major histocompatibility complex (MHC) region (chromosome 6: 25- 34 Mb). We defined significant heritability enrichment as those with an FDR < 0.05.

### Genetic correlation

We estimated the genetic correlations using a bivariate LD score regression (Bulik-Sullivan et al., 2015) using the results from the current GWAS: 99 complex traits (42 diseases and 57 quantitative traits) in Japanese (Akiyama et al., 2017; Ishigaki et al., 2020; Kanai et al., 2018). We excluded variants found in the MHC region from the analysis because of their complex LD structure. We set the significance threshold for genetic correlations as FDR < 0.05. We evaluated the genetic correlation only for ALL-OPLL because the sample sizes of the C- and T-OPLL groups were too small for this analysis.

### MR

We applied two-sample MR methods that handle summary statistics obtained from separate studies to evaluate the causality of BMI, T2D, cerebral aneurysm, and BMD on OPLL using the R package “TwoSampleMR” (Hemani et al., 2018). Regarding BMI, we reconstructed trans-ancestral meta-analysis data using Japanese and European GWAS results in the same way as previously reported (Akiyama et al., 2017; Locke et al., 2015). Regarding T2D, cerebral aneurysm, and BMD, we used publicly available results of East Asian meta-analysis of GWASs for T2D (Spracklen et al., 2020), mainly European meta-analysis of GWAS for cerebral aneurysm (Bakker et al., 2020), and European GWAS for BMD (Kemp et al., 2017). In the SNP selection, we extracted the lead SNPs in the significant independent loci of each study, which were also included in the GWAS meta-analysis for OPLL, and used them as the instrumental variables. When the lead SNP was not present in the GWAS meta-analysis for OPLL, we selected proxy SNPs highly correlated with the original variants (r^2^ > 0.8). If there were no proxy SNPs that met the criteria, we excluded the SNPs from the analysis. The details of the instrument variables in each MR are shown in Supplementary Fig. 9 and Supplementary Table 13.

We conducted additional MR methods in addition to the conventional IVW method for sensitivity analyses: the MR-Egger method (Bowden et al., 2015; Burgess and Thompson, 2017) and the simple and weighted median methods (Bowden et al., 2016). In addition, we conducted subtype-stratified analyses using summary statistics from C- and T-OPLL GWAS and examined the differences between OPLL subtypes. The number of SNPs used in each analysis is listed in Supplementary Table 13. We also assessed the potential bias in the results of MR with leave-one-out analysis and funnel plots.

We performed reverse-direction MR using the lead SNPs in the significant locus in the meta-analysis for ALL-OPLL as instrumental variables. Parts of these OPLL-associated SNPs were not present within the BMI and cerebral aneurysm data sets, although all were contained in the dataset of T2D and BMD. Therefore, in the analysis for BMI and cerebral aneurysm, we substituted them with the proxy SNP in the same way as described above (Supplementary Fig. 9).

A participant overlap between the samples used to estimate genetic associations with the exposure and the outcome in two-sample MR can bias results (Burgess et al., 2016). Therefore, the use of exposure and outcome instrument variables estimated in non-overlapping samples is preferable. We checked the cohort data used in our MR and found that the control samples used in the OPLL study overlapped with up to 2.2% of samples used in the BMI study and 3.4% in the T2D study. According to a simulation study of the association between sample overlap and the degree of bias in instrumental variable analysis, an unbiased estimate is obtained if the overlapping sample includes only control samples for the binary outcome (Burgess et al., 2016).

### Comparison of the effect sizes of the SNPs between the GWAS meta-analyses for OPLL and BMI

After pruning the SNPs, we evaluated the correlations of the effect sizes of the SNPs between the GWAS meta-analyses for OPLL (ALL-, C-, and T-OPLL) and the GWAS for BMI for sets of SNPs stratified by the thresholds based on the GWAS P-values for each trait. We used the results of the GWAS meta-analysis of OPLL and the Japanese GWAS of BMI (Akiyama et al., 2017) for this analysis. First, we extracted SNPs with MAF ≥ 0.01, shared between the meta-analyses for OPLL and BMI. Next, we conducted LD pruning of the SNPs for the SNP pairs in LD (*r*^2^ ≥ 0.5) using 1KGP3 East Asian and JEWEL 3K data by PLINK. Finally, 367,672 SNPs were used in subsequent analyses. We calculated the correlation of the effect sizes of the SNPs between the GWAS meta-analyses for OPLL and BMI GWAS for sets of SNPs stratified by the thresholds based on the GWAS P-values in each trait using R statistical software version 3.6.3.

### Generation of PRS of BMI and its application to OPLL GWAS samples

We used PRS to investigate the genetic impact of BMI on OPLL. We constructed the PRS of BMI using a pruning and thresholding method (Khera et al., 2018) (Supplementary Fig. 19). In the discovery phase, we generated PRS as the sum of risk alleles weighted by the log odds ratio of association estimated in the Japanese GWAS for BMI (Akiyama et al., 2017). We pruned SNPs based on nine different LD thresholds r^2^ = 0.1- 0.9 in a 250-kb window using PLINK2.0 and constructed 20 PRS using independent SNPs at P-value thresholds of 5.0 × 10^-8^ ∼ 1 for each LD threshold. In the validation phase, we determined the best pruning parameter with another Japanese dataset in which genotyping was conducted using the Illumina OmniExpressExome chip. We conducted data quality control in the same manner as in the OPLL GWAS. We calculated the Spearman’s rho score between BMI and PRS to assess the fit of the models and determined r^2^ = 0.9 and P-value = 0.6 as the best parameter. We applied BMI PRS for cases and controls in the present OPLL GWAS study in the test phase.

We measured the association between BMI PRS and OPLL using logistic regression with principal components 1-10 as covariates for each OPLL dataset. Then, we meta-analyzed the effect sizes of BMI PRS in the three OPLL datasets using the inverse-variance method under a fixed-effect model. We conducted this analysis for ALL-OPLL, C-, and T-OPLL. Furthermore, we applied BMI PRS for C- and T-OPLL cases and compared the effect of OPLL PRS on OPLL subtypes using logistic regression with principal components 1-10 as covariates.

### Data availability

Full GWAS results will be available after acceptance via the website of the Japanese ENcyclopedia of GEnetic associations by Riken (JENGER, http://jenger.riken.jp/en/).

## Supporting information

Supplementary Figures, Notes, and references

## Acknowledgments

We thank the OPLL patients and their families who participated in this study and the OPLL patient network in Japan (Zensekichuren. President, Yasuko Masuda). This work was supported by a JSPS KAKENHI Grant (no. 19K09566 to T.E.), a grant from Japan Orthopaedics and Traumatology Foundation (no. 452 to Y. Koike), a grant from AO Spine Japan Research (to M. Takahata), a grant from Suhara Memorial Foundation (to M. Takahata), grants from Japan Agency for Medical Research and Development (AMED) (no. JP21kk0305013, JP21tm0424220, and JP21ck0106642 to C.T.), and the JCR Grant for Promoting Basic Rheumatology (to C.T.).

## Author contributions

Conceptualization: S Ikegawa, M Takahata and CT Data curation: YI, SN, KO and YM

Formal analysis: Y Koike, MN, NO, H Suetsugu, XL, S Ito, K Tomizuka, KH and CT Methodology: S Ikegawa and CT

Visualization: Y Koike

Funding acquisition: Y Koike, M Takahata, TE and CT Project administration: S Ikegawa, M Takahata and CT

Resources: Y Koike, M Takahata, MN, TE, S Imagama, K Kobayashi, T Kaito, S Kato, Y Kawaguchi, MK, H Sakai, T Tsuji, TM, HI, TY, MK, H Nakashima, KA, YT, K Takeuchi, SM, H Nakajima, KM, AS, SF, T Kanchiku, KW, T Tanaka, K Kida, S Kobayashi, M Tahahashi and KY Supervision: Y Kamatani, MY, AO, MM and NI

Writing – original draft: Y Koike, MN, HT and HFL Writing – review & editing: M Takahata, CT and S Ikegawa

## Notes

### Competing Interest Statement

Yoshiharu Kawaguchi: Consulting fees from Medacta International, Masashi Yamazaki: A representative of Japanese Organization of the Study for Ossification of Spinal Ligament, Atsushi Okawa: A representative of Japanese Organization of the Study for Ossification of Spinal Ligament, The other authors declare that no competing interests exist.

### Author Declarations

This research project was approved by the ethical committees of RIKEN Centers for Integrative Medical Sciences, Hokkaido University, Keio University School of Medicine, Nagoya University, Osaka University, Kanazawa University, the University of Toyama, Hakodate Central General Hospital, Spinal Injuries Center, Tokyo Medical and Dental University, the University of Tokyo, National Okayama Medical Center, Kagoshima University, the University of Fukui, Shiga University of Medical Science, Jichi Medical University, Kyoto University, Yamaguchi University, Niigata University, Hirosaki University, Kochi Medical School, Hamamatsu University School of Medicine, Kyorin University School of Medicine, Kurume University School of Medicine, and National Center for Geriatrics and Gerontology.

