## Supplementary Figures, Notes, and references for "Genetic insights into ossification of the posterior longitudinal ligament of the spine"

### Supplementary Information

#### Supplementary Figures:


**Supplementary Fig. 1**. The overview of the genome-wide association study meta-analysis

**Supplementary Fig. 2**. A quantile-quantile plot of meta-analysis of genome-wide association studies

**Supplementary Fig. 3**. Regional association plots for 14 susceptibility loci for OPLL

**Supplementary Fig. 4**. Summary-data-based Mendelian randomization

**Supplementary Fig. 5**. OPLL-subtype stratification identified subtype-specific loci

**Supplementary Fig. 6** | A quantile-quantile plot of meta-analysis of subtype stratified genome-wideassociation studies

**Supplementary Fig. 7**. Regional association plots for 3 susceptibility loci for cervical OPLL

**Supplementary Fig. 8**. Regional association plots for 8 susceptibility loci for thoracic OPLL

**Supplementary Fig. 9.** Selection of SNPs to be used as instrumental variables in Mendelian randomization

**Supplementary Fig. 10**. Scatter plots for the Mendelian randomization of the causal effect of BMI on OPLL

**Supplementary Fig. 11**. Sensitivity analysis of the Mendelian randomization of BMI causality on OPLL

**Supplementary Fig. 12**. Scatter plots for the Mendelian randomization of the causal effect of type 2 diabetes on OPLL

**Supplementary Fig. 13**. Scatter plots for the Mendelian randomization of the causal effect of bone mineral density on OPLL

**Supplementary Fig. 14**. Scatter plots for the Mendelian randomization of the causal effect of cerebral aneurysm on OPLL

**Supplementary Fig. 15**. Scatter plots for the Mendelian randomization of the causal effect of OPLL on BMI, type 2 diabetes, cerebral aneurysm, and bone mineral density

**Supplementary Fig. 16**. Scatter plots for the Mendelian randomization of the causal effect of BMI on OPLL subtypes

**Supplementary Fig. 17**. Sensitivity analysis of the Mendelian randomization of BMI causality on OPLL Subtypes

**Supplementary Fig. 18**. Correlation of the effect sizes of the genome-wide SNPs of OPLL and BMI

**Supplementary Fig. 19**. BMI polygenic risk score analysis for OPLL

**Supplementary Fig. 20**. Determination of the best parameter for BMI polygenic risk score

**Supplementary Notes:**

**Supplementary Note 1**. Members of Genetic Study Group of Investigation Committee on Ossification of the Spinal Ligaments

**Supplementary Note 2**. SNP-obesity interaction

**Supplementary reference**

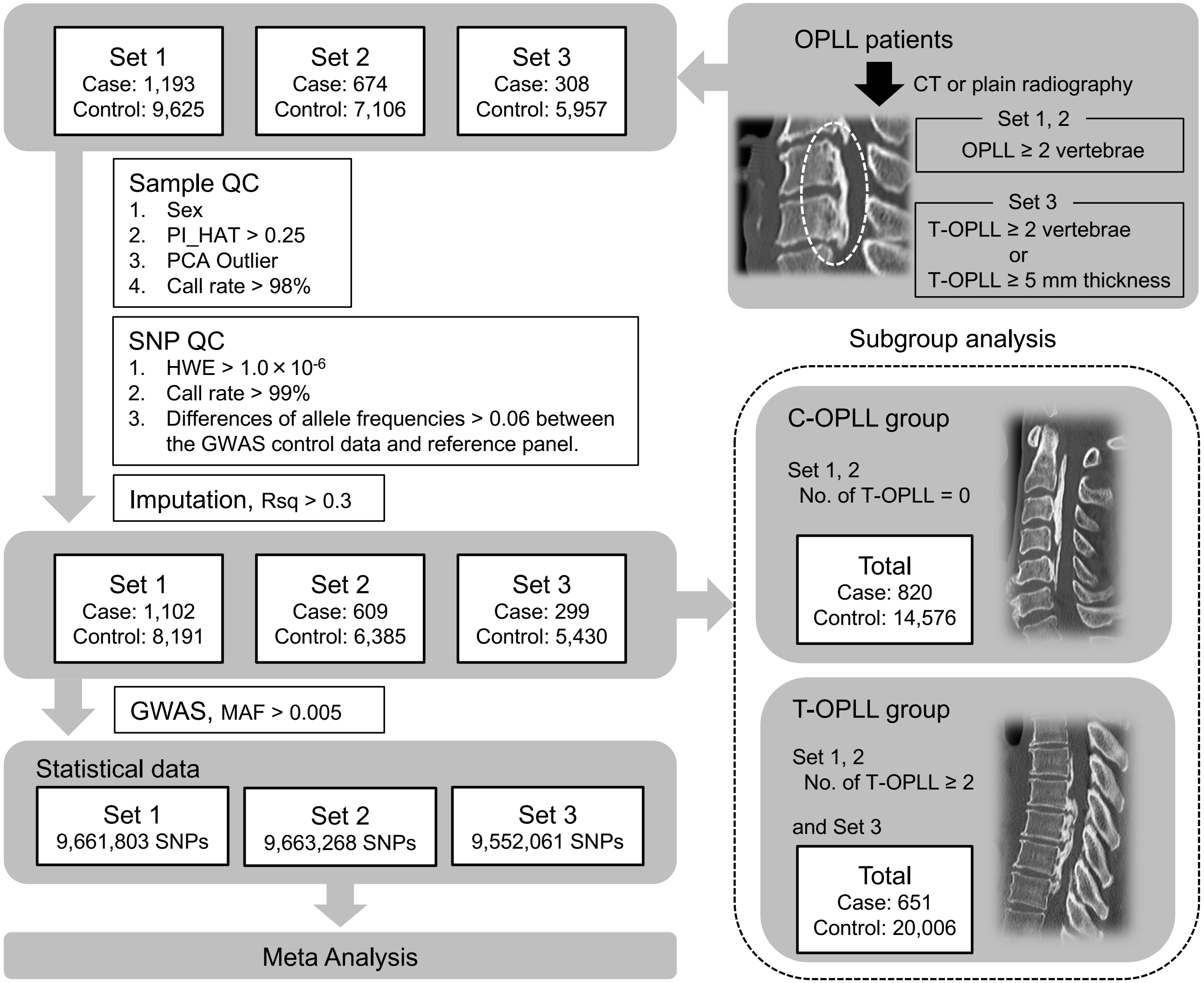


**Supplementary Fig. 1 | The overview of the genome-wide association study meta-analysis**

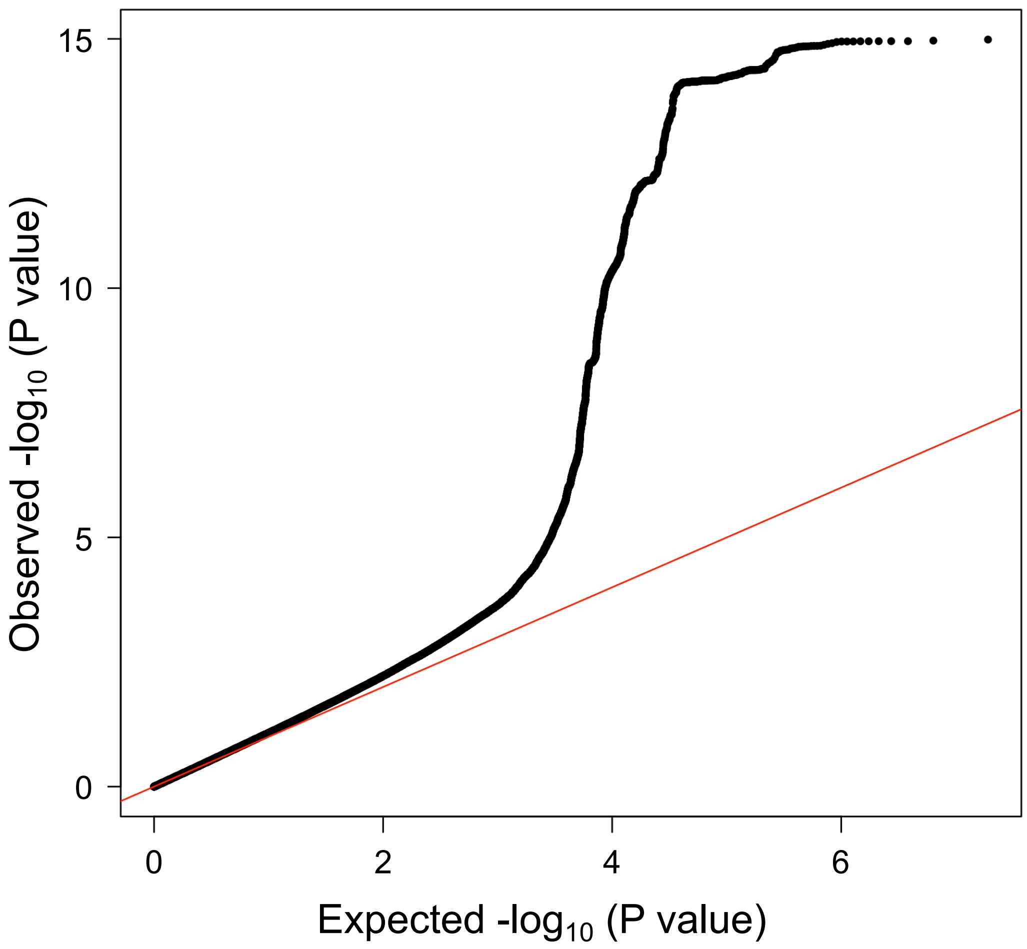


**Supplementary Fig. 2 | A quantile-quantile plot of meta-analysis of genome-wide association studies**

Horizontal and vertical lines represent the expected P-value under a null distribution and the observed P-value, respectively.

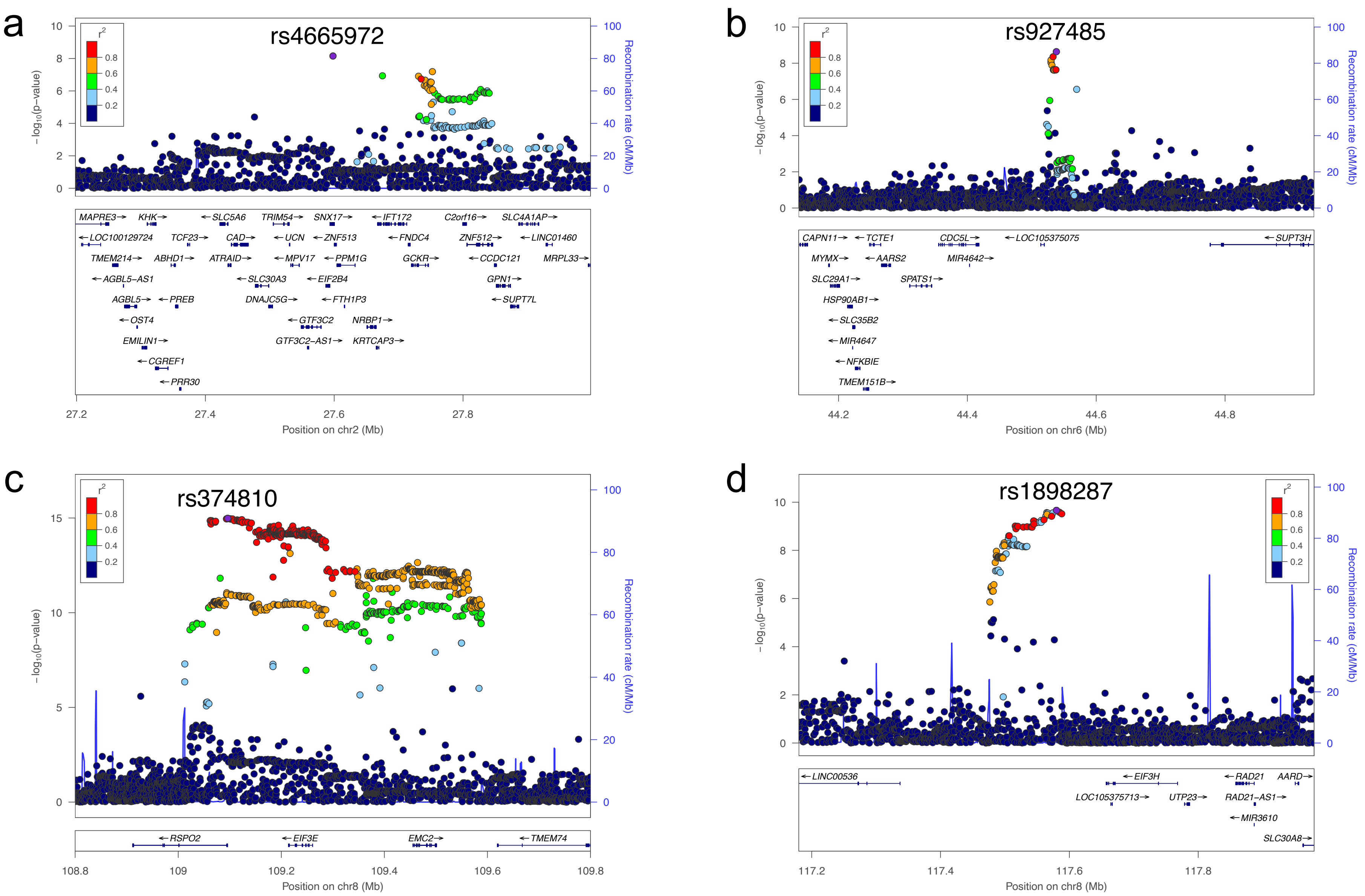
a
rs4665972
b
rs927485
c
rs374810
d
rs1898287
−log10(p−value)
Recombination rate (cM/Mb)
r2
Position on chr2 (Mb)
Position on chr6 (Mb)
Position on chr8 (Mb)
Position on chr8 (Mb)
MAPRE3 KHK SLC5A6 TRIM54 SNX17 IFT172 C2orf16 SLC4A1AP
LOC100129724 TCF23 CAD UCN ZNF513 FNDC4 ZNF512 LINC01460
TMEM214 ABHD1 ATRAID MPV17 PPM1G GCKR CCDC121 MRPL33
AGBL5−AS1 SLC30A3 EIF2B4 GPN1
AGBL5 PREB DNAJC5G FTH1P3 SUPT7L
OST4 GTF3C2 NRBP1
EMILIN1 GTF3C2−AS1 KRTCAP3
CGREF1
PRR30
CAPN11 TCTE1 CDC5L LOC105375075 SUPT3H
MYMX AARS2 MIR4642
SLC29A1 SPATS1
HSP90AB1
SLC35B2
MIR4647
NFKBIE
TMEM151B
RSPO2 EIF3E EMC2 TMEM74
LINC00536 EIF3H RAD21 AARD
LOC105375713 UTP23 RAD21−AS1
MIR3610
SLC30A8

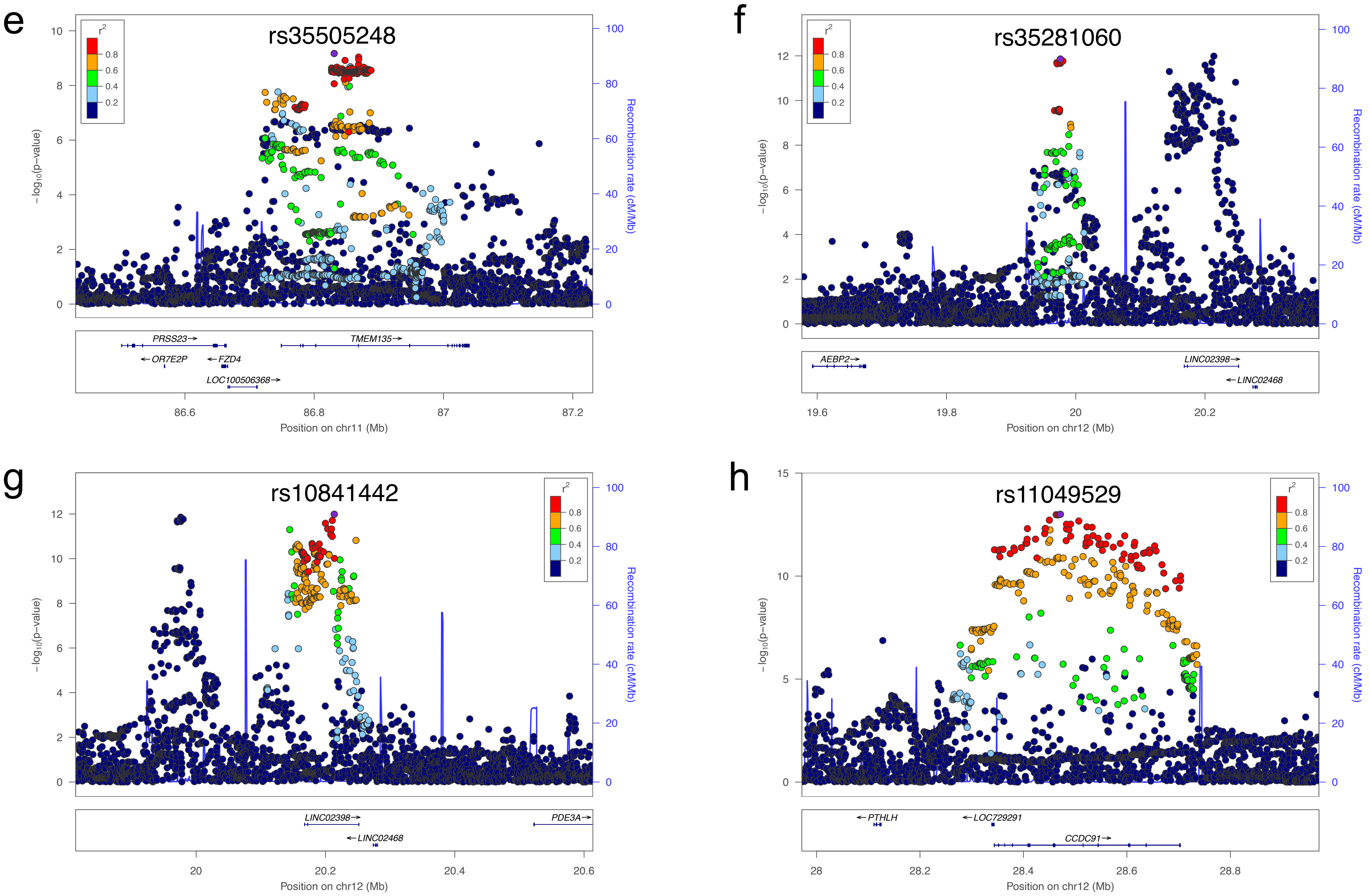
e
rs35505248
r2
0.8
0.6
0.4
0.2
−log10(p−value)
Recombination rate (cM/Mb)
PRSS23→
←OR7E2P
←FZD4
LOC100506368→
TMEM135→
86.6
86.8
87
87.2
Position on chr11 (Mb)
f
rs35281060
AEBP2→
LINC02398→
←LINC02468
19.6
19.8
20
20.2
Position on chr12 (Mb)
g
rs10841442
LINC02398→
←LINC02468
PDE3A→
20
20.2
20.4
20.6
Position on chr12 (Mb)
h
rs11049529
←PTHLH
←LOC729291
CCDC91→
28
28.2
28.4
28.6
28.8
Position on chr12 (Mb)

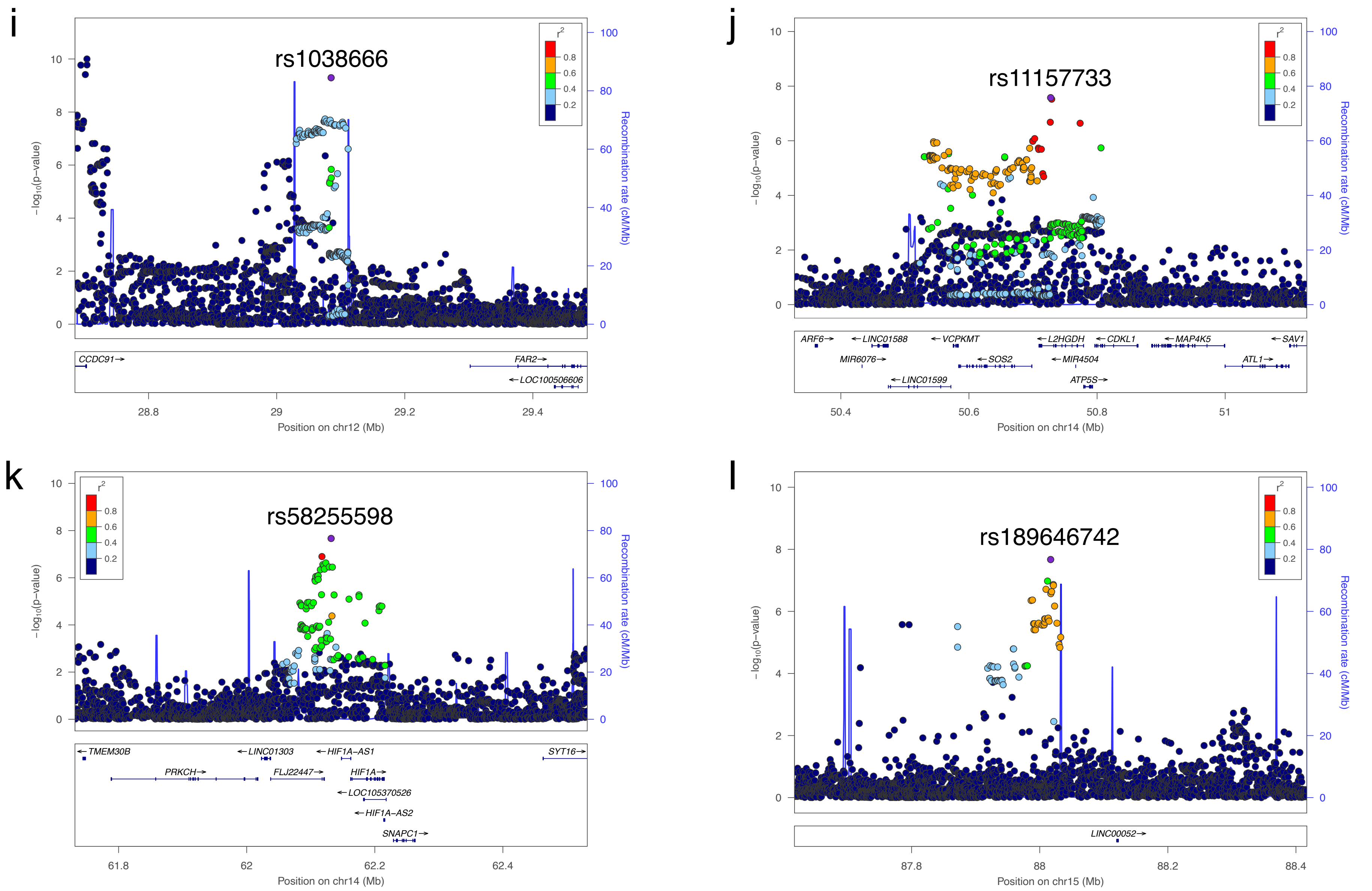
i
rs1038666
r2
0.8
0.6
0.4
0.2
-log10(p-value)
Recombination rate (cM/Mb)
CCDC91→
FAR2→
←LOC100506606
28.8
29
29.2
29.4
Position on chr12 (Mb)
j
rs11157733
ARF6→
←LINC01588
←VCPKMT
←L2HGDH
←CDKL1
←MAP4K5
←SAV1
MIR6076→
←SOS2
←MIR4504
ATL1→
←LINC01599
ATP5S→
50.4
50.6
50.8
51
Position on chr14 (Mb)
k
rs58255598
←TMEM30B
←LINC01303
←HIF1A-AS1
SYT16→
PRKCH→
FLJ22447→
HIF1A→
←LOC105370526
←HIF1A-AS2
SNAPC1→
61.8
62
62.2
62.4
Position on chr14 (Mb)
l
rs189646742
LINC00052→
87.8
88
88.2
88.4
Position on chr15 (Mb)

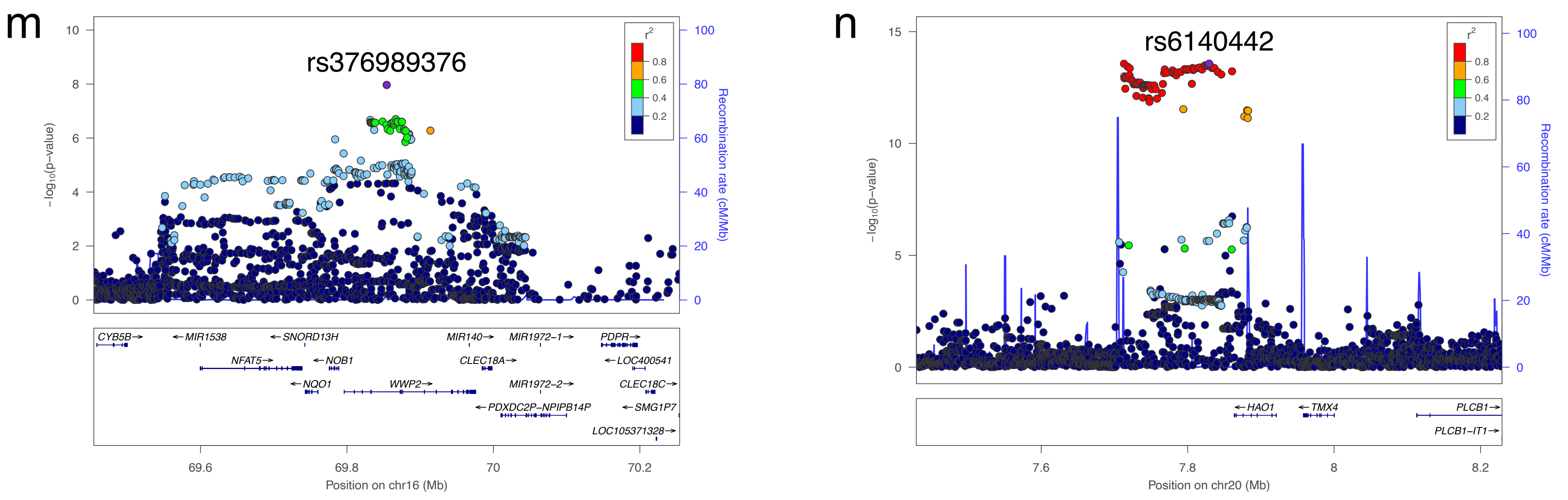


**Supplementary Fig. 3 | Regional association plots for 14 susceptibility loci for OPLL**

Each plot shows $-\log_{10}$ P-values against the chromosomal position of variants in specific regions. (a) 2p23.3. (b) 6p21.1. (c) 8q23.1. (d) 8q23.3. (e) 11q14.2 (f) 12p12.3. (g) 12p12.2. (h) 12p11.22. (i) 12p11.22. (j) 14q21.3. (k) 14q23.2. (l) 15q25.3. (m) 16q22.1. (n) 20p12.3. The variant with the highest association signal in each locus is represented in purple; the other variants are colored according to the extent of LD with this variant. The estimated recombination rates from hg19/1000 Genomes Nov 2014 East Asian are shown as light blue lines.

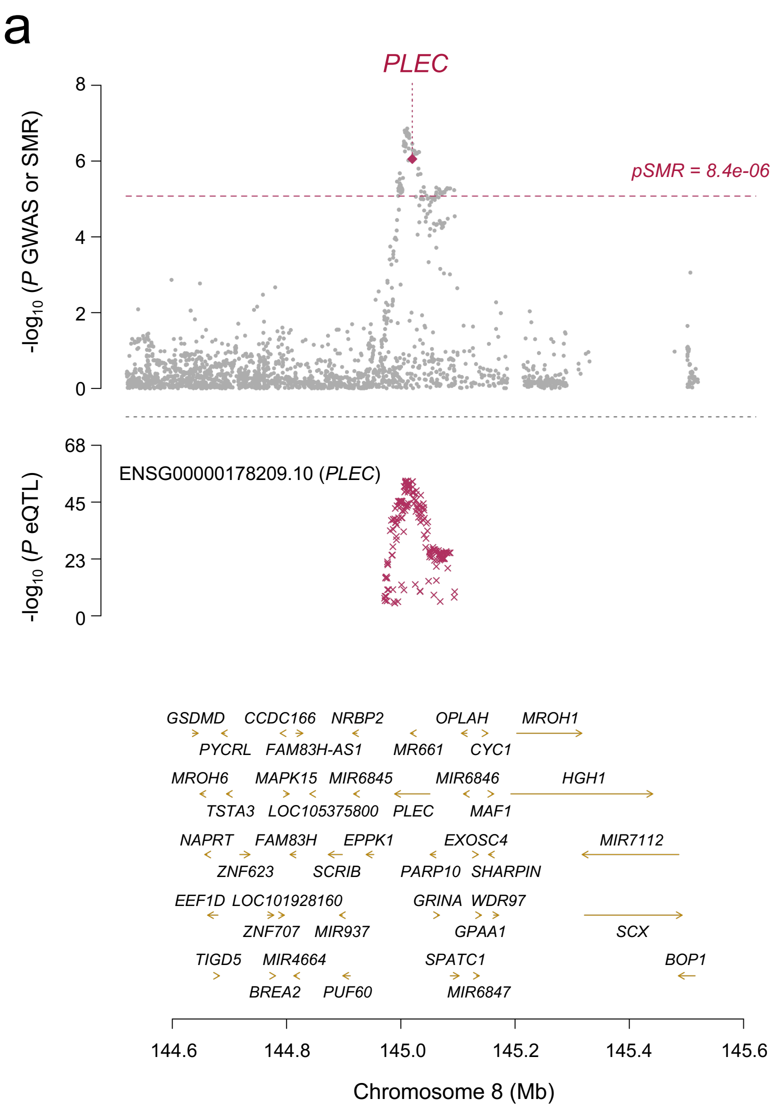


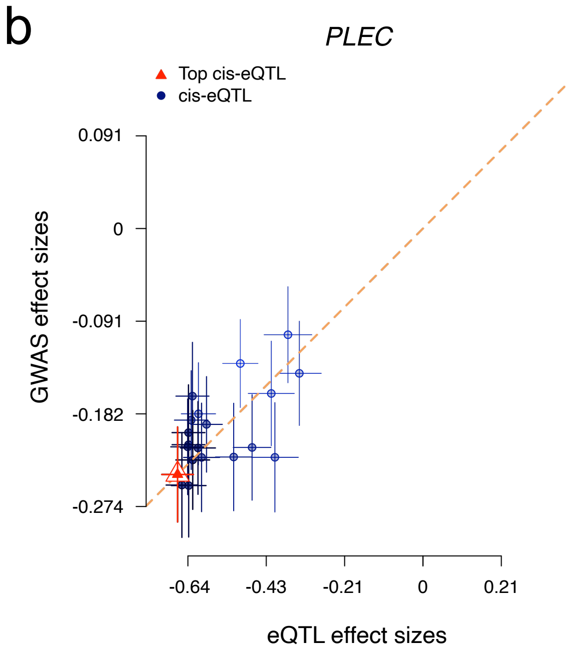


**Supplementary Fig. 4 | Summary-data-based Mendelian randomization**

(a) In the top plot, gray dots represent P-values for SNPs from the GWAS meta-analysis for OPLL, and diamonds represent P-values for probes from the SMR. In the bottom plot, each red “X” represents the eQTL P-value of SNPs from GTEx v7 for PLEC in fibroblast. eQTL, expression quantitative trait locus; GTEx, Genotype-Tissue Expression.

(b) Effect sizes of SNPs from GWAS plotted against those for SNPs from the fibroblast eQTL study. Orange dashed line represents the estimate of the effect size of the SMR at the top cis-eQTL. Error bars are standard errors of SNP effects.

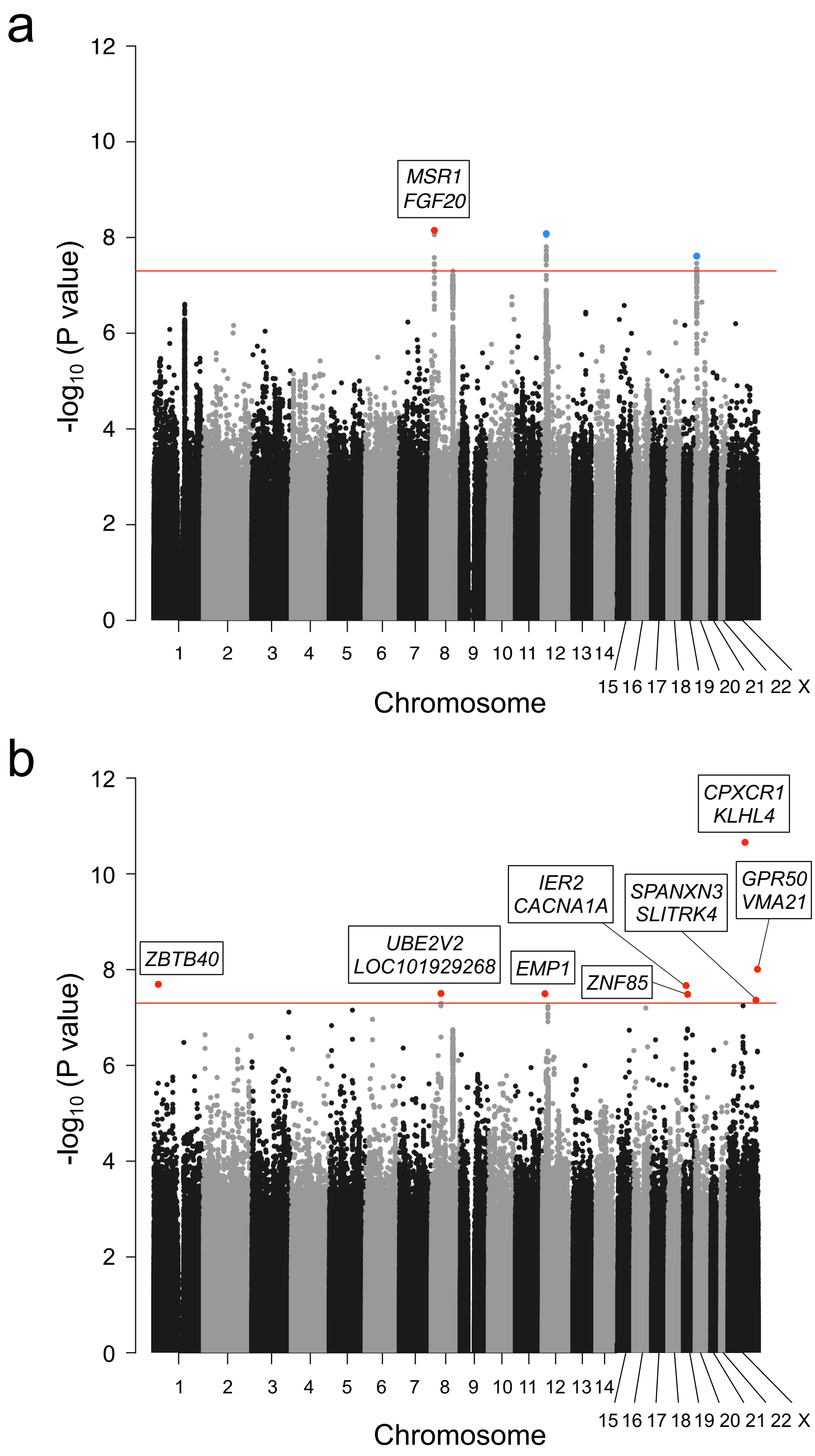


**Supplementary Fig. 5 | OPLL-subtype stratification identified subtype-specific loci**

Manhattan plot showing the -$\log_{10}$ P-value for each SNP in the GWAS meta-analysis. (a) Cervical OPLL (b) Thoracic OPLL The values were plotted against the respective chromosomal positions. The horizontal red line represents the genome-wide significance threshold ($P = 5.0 \times 10^{-8}$). Red and blue points represent the lead SNPs in the new and known loci, respectively.

a

b

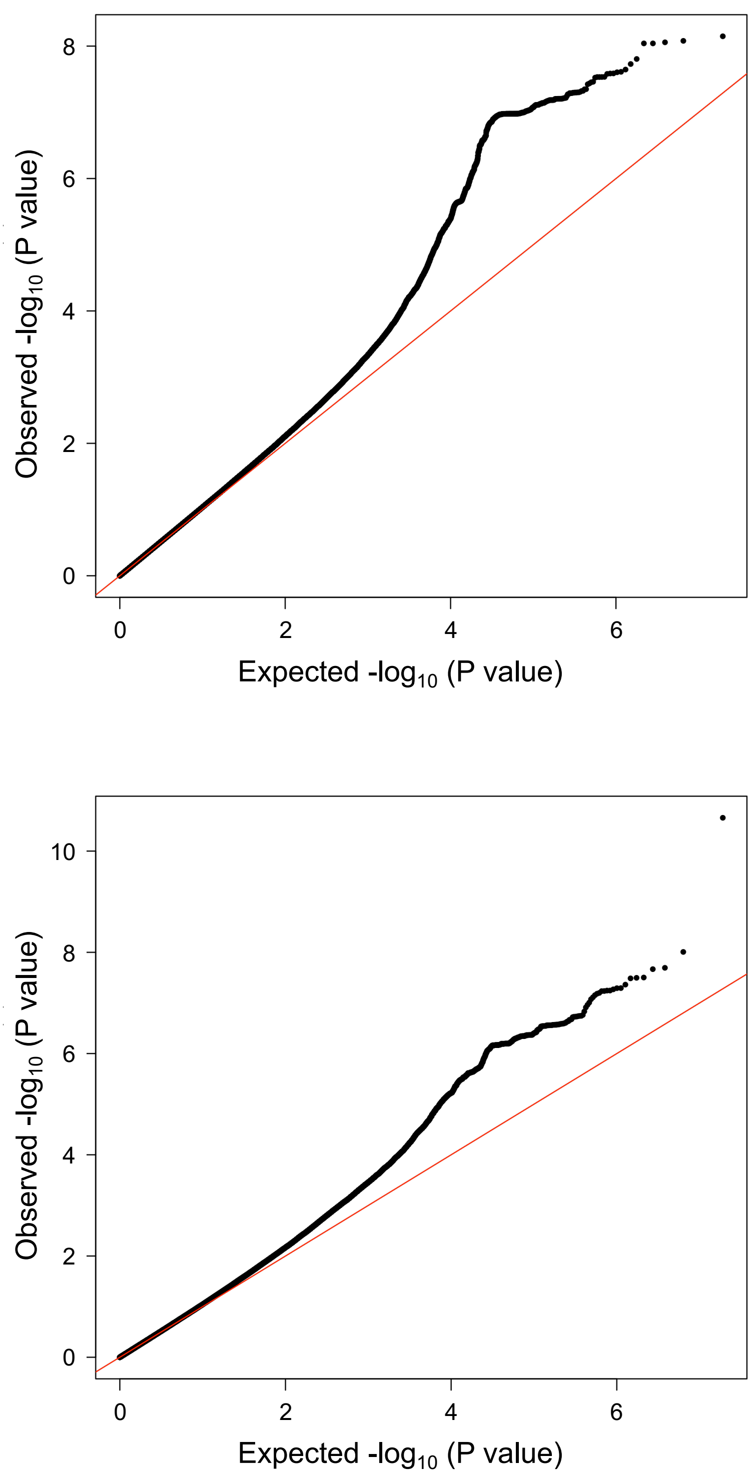


**Supplementary Fig. 6 | A quantile-quantile plot of meta-analysis of subtype stratified genome-wide association studies**

Horizontal and vertical lines represent the expected P-value under a null distribution and the observed P-value, respectively. (a) Cervical OPLL (b) Thoracic OPLL

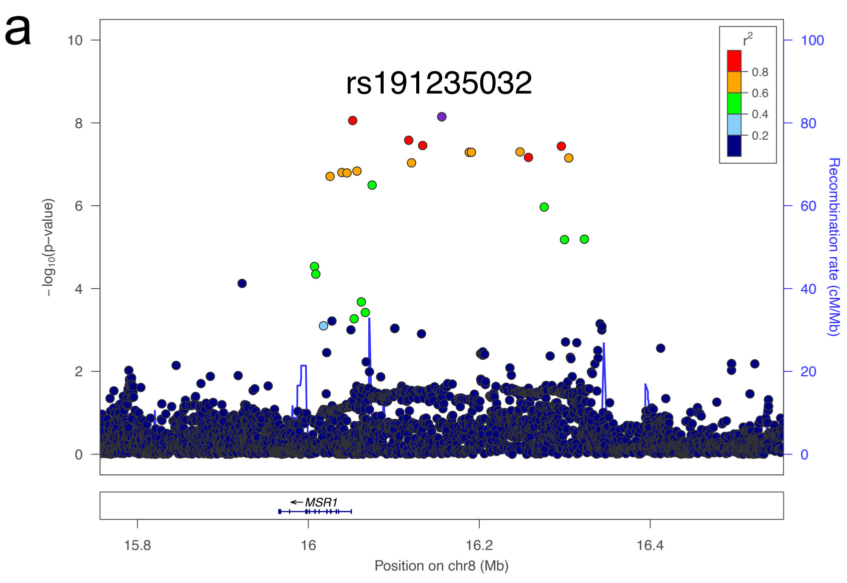


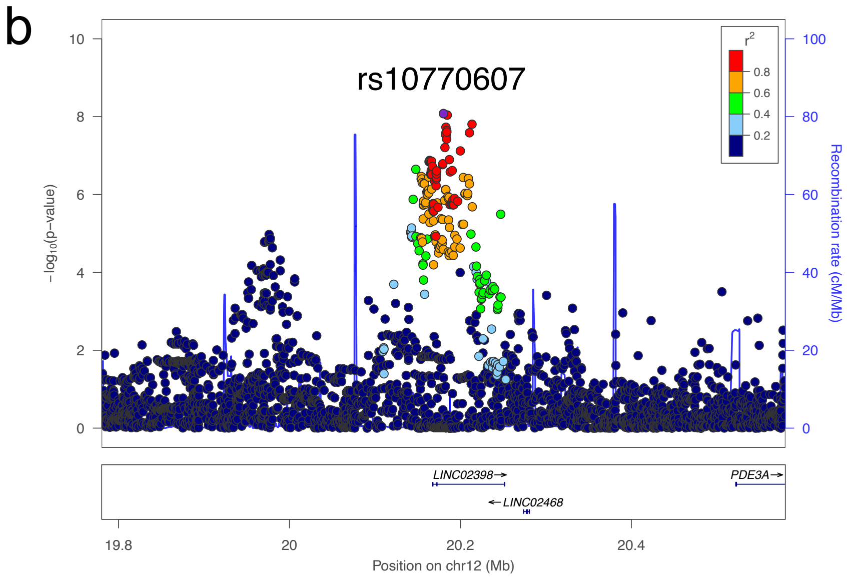


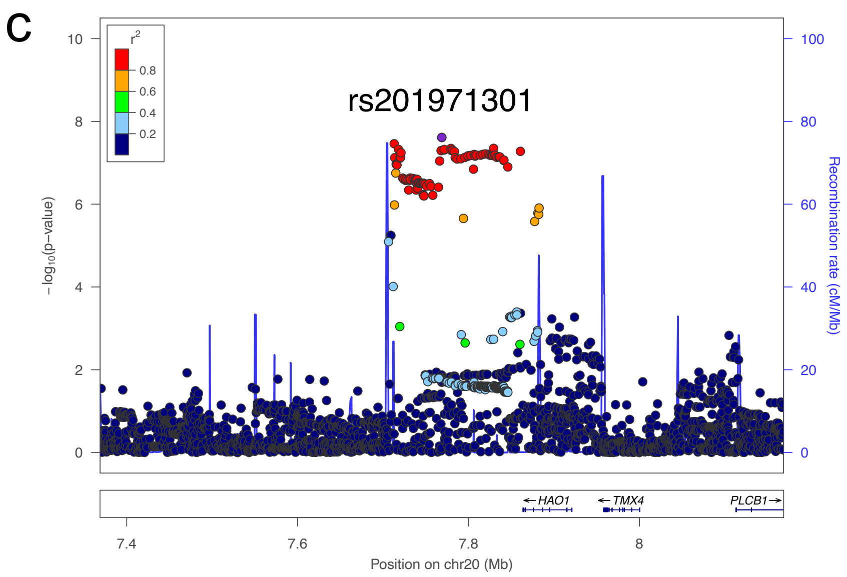


**Supplementary Fig. 7 | Regional association plots for three susceptibility loci for cervical OPLL**

Each plot shows $-\log_{10}$ P values against the chromosomal position of variants in a specific region. (a) 8p22. (b) 12p12.2. (c) 20p12.3. The variant with the highest association signal in each locus is represented in purple; the other variants are colored according to the extent of LD with this variant. The estimated recombination rates from hg19/1000 Genomes Nov 2014 East Asian are shown as light blue lines.

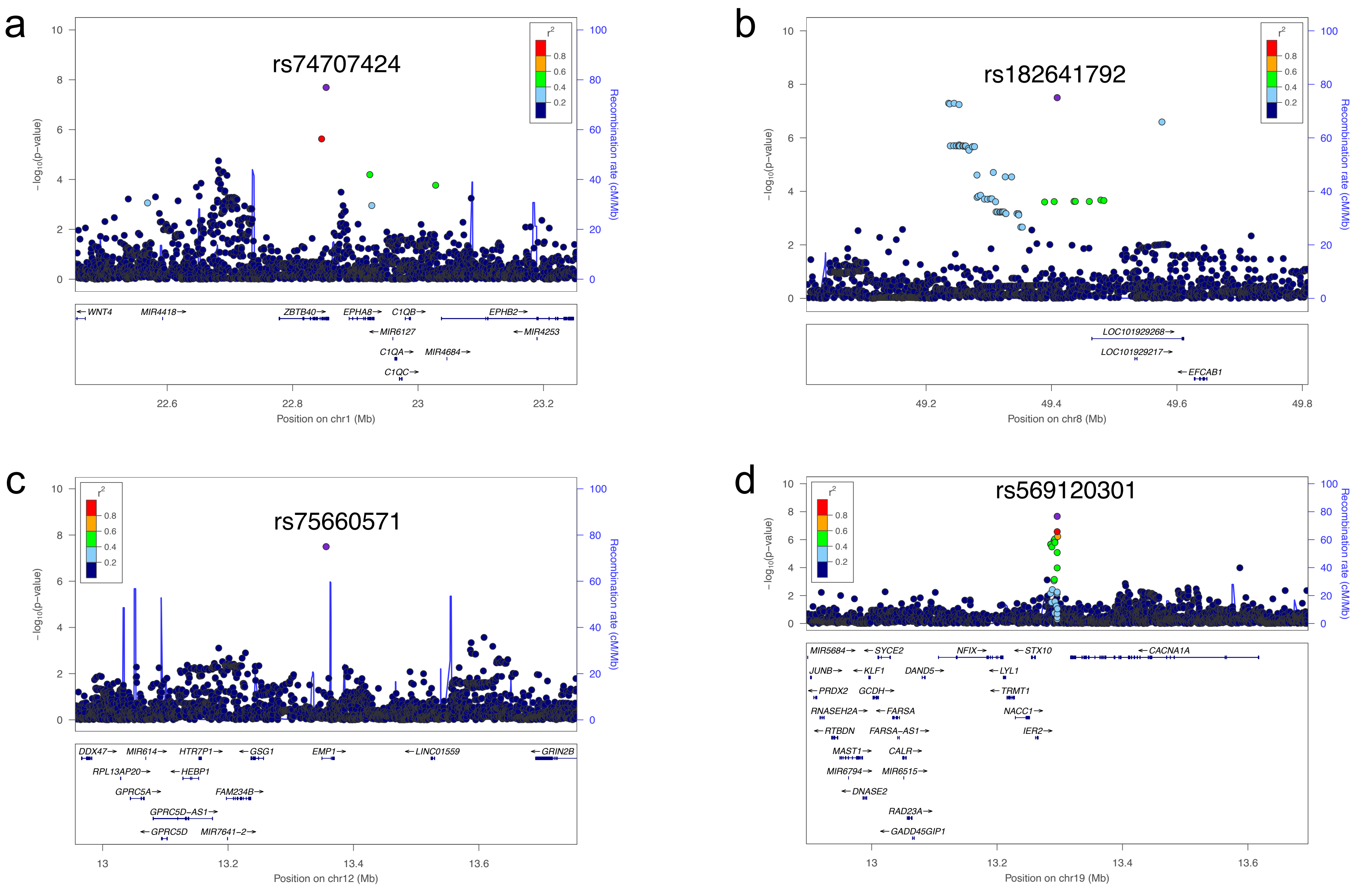

a
rs74707424
b
rs182641792
c
rs75660571
d
rs569120301
−log10(p−value)
Recombination rate (cM/Mb)
r2
Position on chr1 (Mb)
Position on chr8 (Mb)
Position on chr12 (Mb)
Position on chr19 (Mb)
WNT4
MIR4418
ZBTB40
EPHA8
C1QB
EPHB2
MIR6127
MIR4253
C1QA
MIR4684
C1QC
LOC101929268
LOC101929217
EFCAB1
DDX47
MIR614
HTR7P1
GSG1
EMP1
LINC01559
GRIN2B
RPL13AP20
HEBP1
GPRC5A
FAM234B
GPRC5D−AS1
GPRC5D
MIR7641−2
MIR5684
SYCE2
NFIX
STX10
CACNA1A
JUNB
KLF1
DAND5
LYL1
PRDX2
GCDH
TRMT1
RNASEH2A
FARSA
NACC1
RTBDN
FARSA−AS1
IER2
MAST1
CALR
MIR6794
MIR6515
DNASE2
RAD23A
GADD45GIP1

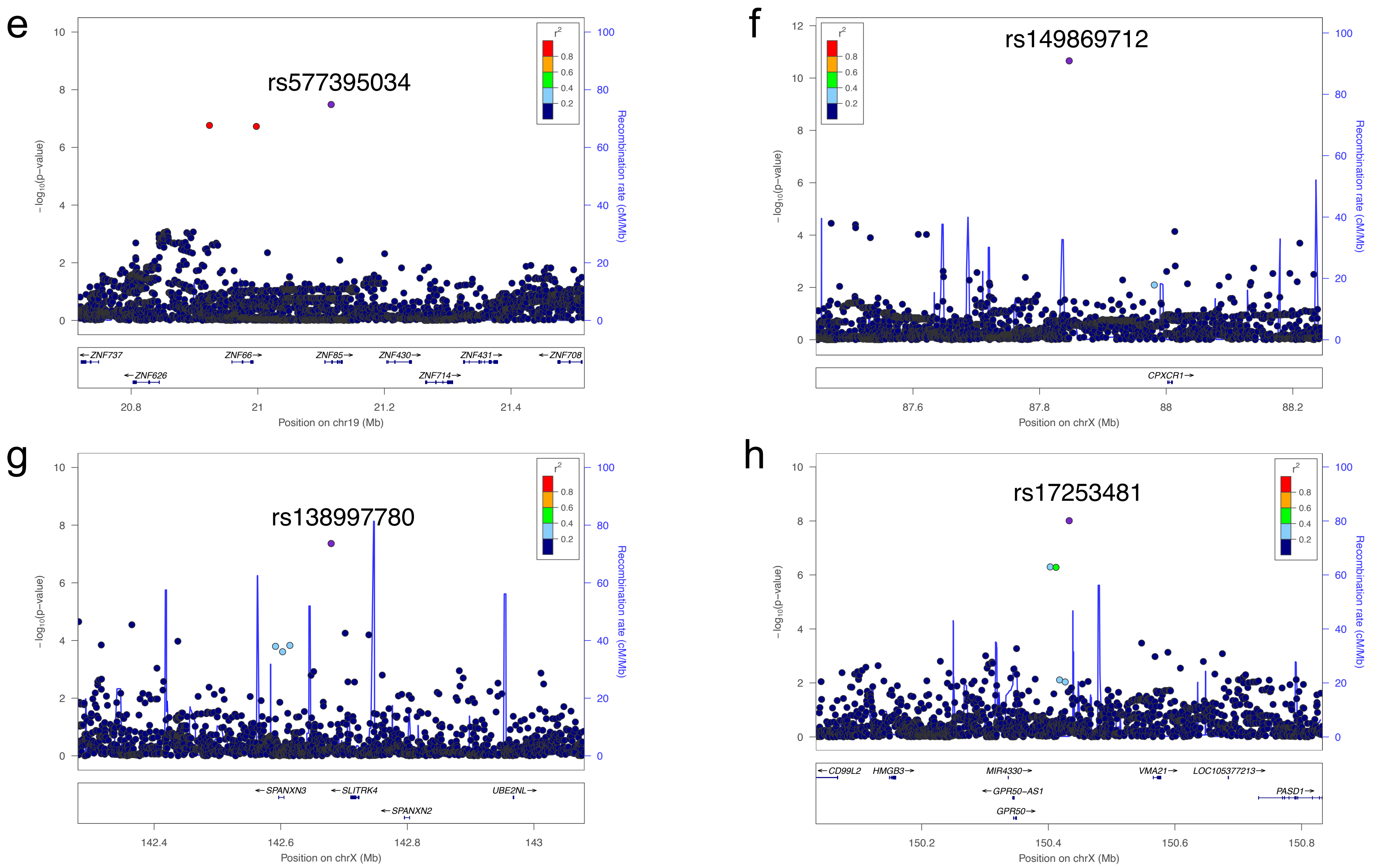


**Supplementary Fig. 8 | See next page for caption**

**Supplementary Fig. 8 | Regional association plots for eight susceptibility loci for thoracic OPLL**

Each plot shows $-\log_{10}$ P values against the chromosomal position of variants in a specific region. (a) 1p36.12. (b) 8q11.21. (c) 12p13.1. (d) 19p13.2. (e) 19p12. (f) 23q23.31. (g) 23q27.3. (h) 23q28. The variant with the highest association signal in each locus is represented in purple; the other variants are colored according to the extent of LD with this variant. The estimated recombination rates from hg19/1000 Genomes Nov 2014 East Asian are shown as light blue lines.

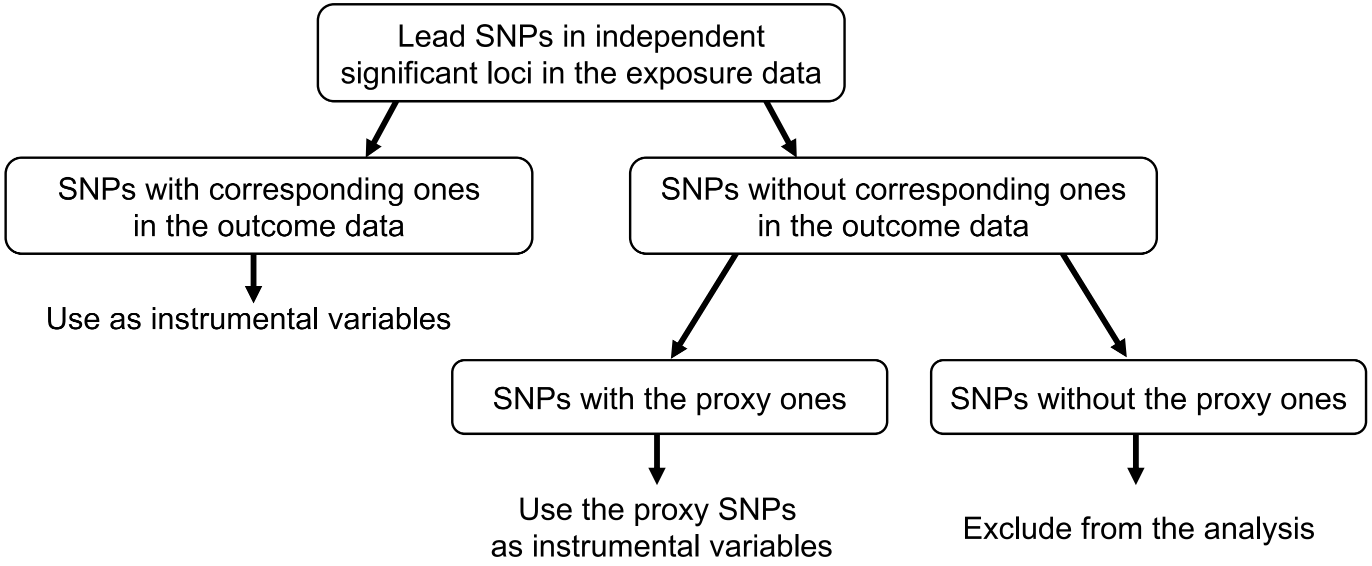


**Selection criteria for the proxy SNP**

1. Strong LD* with the original SNP
2. P-value is lowest and less than $5 \times 10^{-8}$ in the exposure data

*** LD calculation**

| Exposure trait | Data used for calculating LD | Criteria for strong LD |
|---|---|---|
| BMI, cerebral aneurysm, BMD, OPLL | 1KGP3 EAS, 1KGP3 EUR | $r^2$ > 0.8 in both EAS and EUR |
| T2D | 1KGP3 EAS | $r^2$ > 0.8 in EAS |

**Supplementary Fig. 9 | Selection of SNPs to be used as instrumental variables in Mendelian randomization**

1KGP3, the 1000 Genomes Project Phase 3; EAS, East Asian; EUR, European.

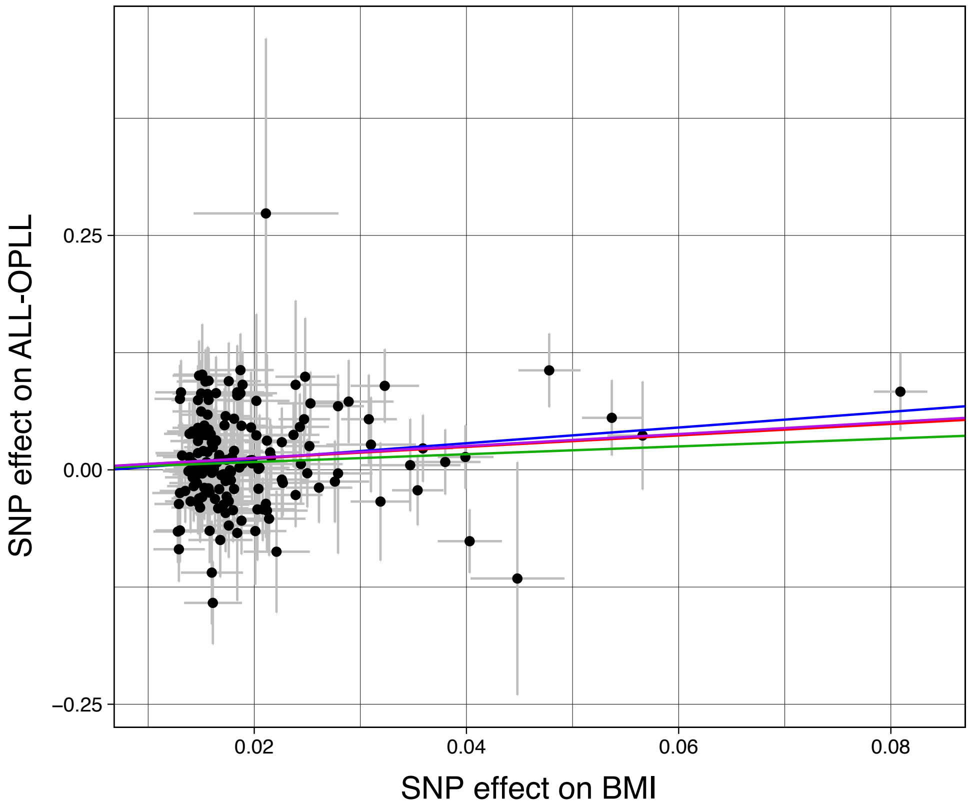


**Supplementary Fig. 10 | Scatter plots for the Mendelian randomization of the causal effect of BMI on OPLL**

Each dot represents the BMI-associated SNP plotted along with the effect size estimates on BMI (x-axis) and OPLL (y-axis). Error bars represent 95% confidence intervals. The slopes of the lines represent the causal association evaluated by four Mendelian randomization (MR) methods: red, inverse variance weighted (IVW); blue, MR-Egger, green, simple median; purple, weighted median.

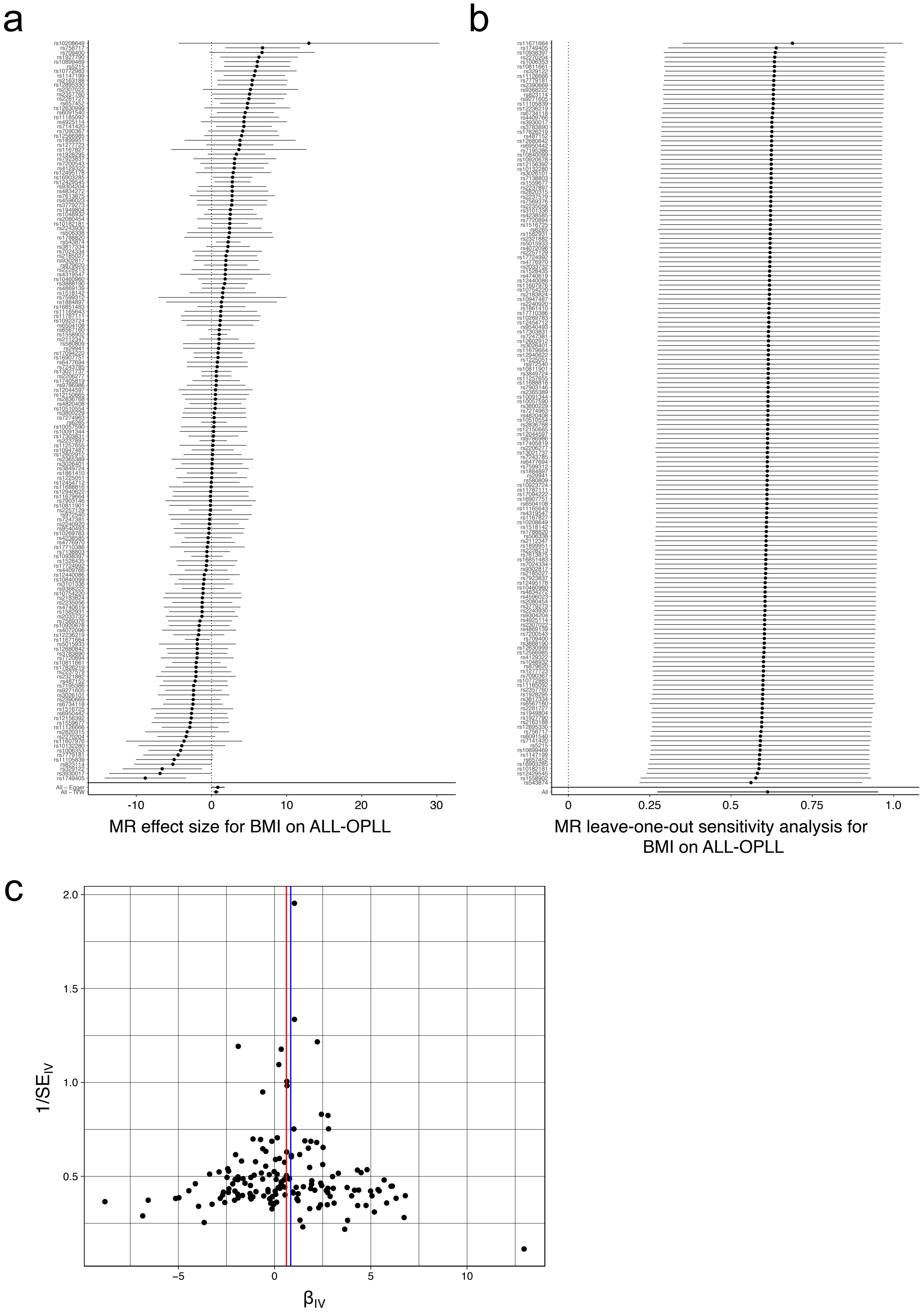


**Supplementary Fig. 11 | See next page for caption**

**Supplementary Fig. 11 | Sensitivity analysis of the Mendelian randomization of BMI causality on OPLL**

(a) Forest plot. Each black point represents an effect size for BMI on OPLL, produced using significant SNPs in BMI GWAS as separate instruments. The black point in the bottom row shows the combined causal estimate using all SNPs together in a single instrument, using two methods of Mendelian randomization (MR): inverse-variance weighted（IVW） and MR-Egger. Horizontal lines are 95% confidence intervals.

(b) Leave-one-out analysis. Each row represents an MR result (IVW) of BMI on ALL-OPLL after discarding the SNP listed on the y-axis. The point represents the effect size, and the horizontal line represents the 95% confidence interval.

(c) Funnel plot. On the y-axis, $1/SE_{IV}$ represents the inverse standard error of the estimated causal effect for each single SNPs (instrumental variables). On the x-axis, $\beta_{IV}$ represents the effect size of each SNP. Colored lines represent the effect sizes of the different MR analyses: red, IVW; blue, MR-Egger.

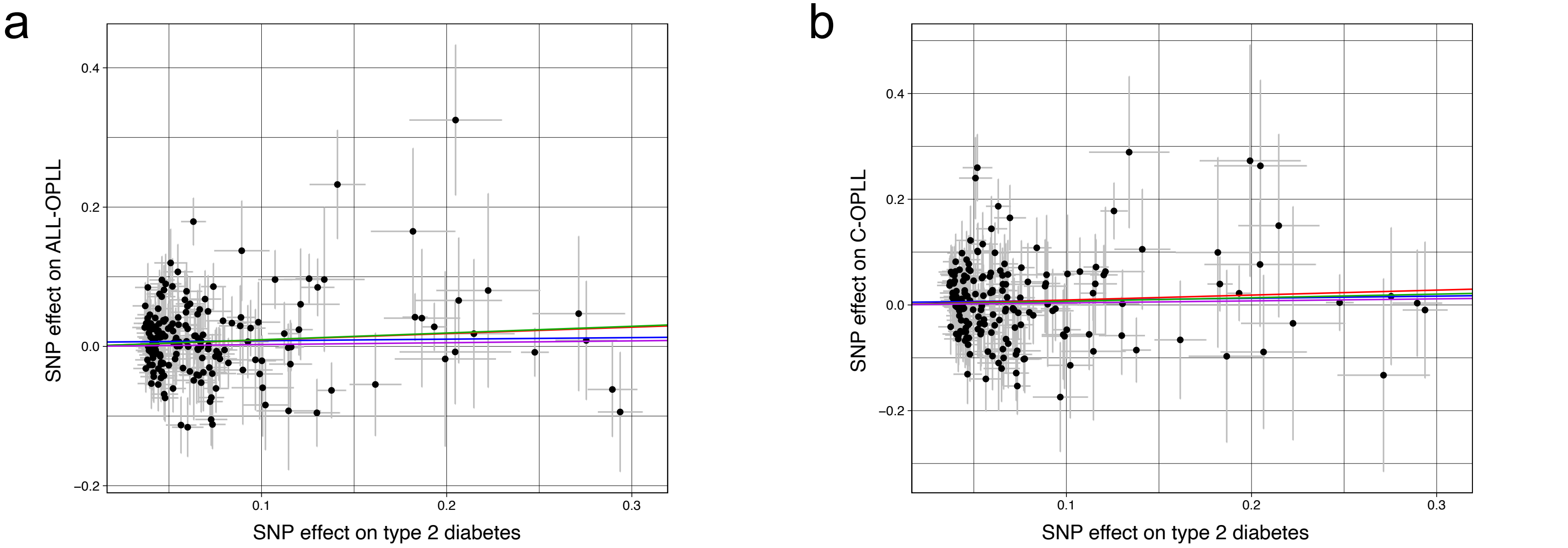


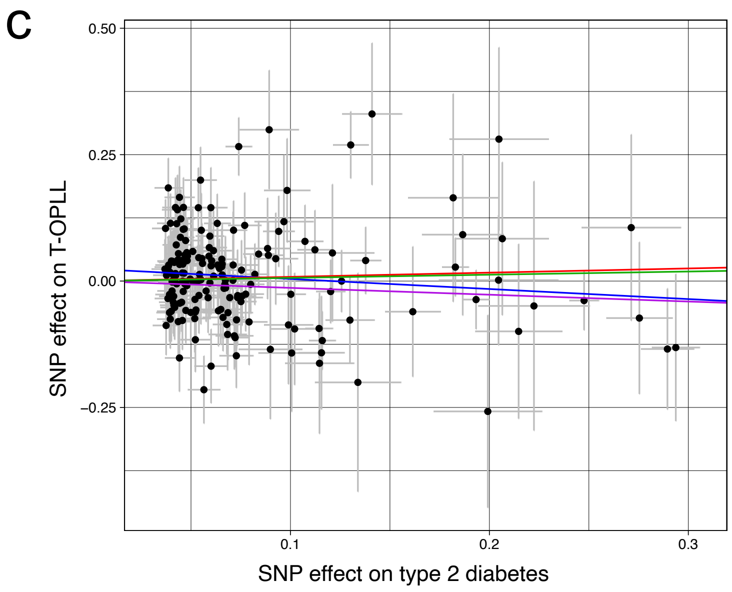


**Supplementary Fig. 12 | Scatter plots for the Mendelian randomization of the causal effect of type 2 diabetes on OPLL**

(a) OPLL. (b) cervical OPLL. (c) thoracic OPLL. Each dot represents the type 2 diabetes-associated SNP plotted along with effect size estimates on type 2 diabetes (x-axis) and OPLL (y-axis). Error bars represent 95% confidence intervals. The slopes of the line represent the causal association evaluated by four Mendelian randomization (MR) methods: red, inverse variance weighted (IVW); blue, MR-Egger; green, simple median; purple, weighted median

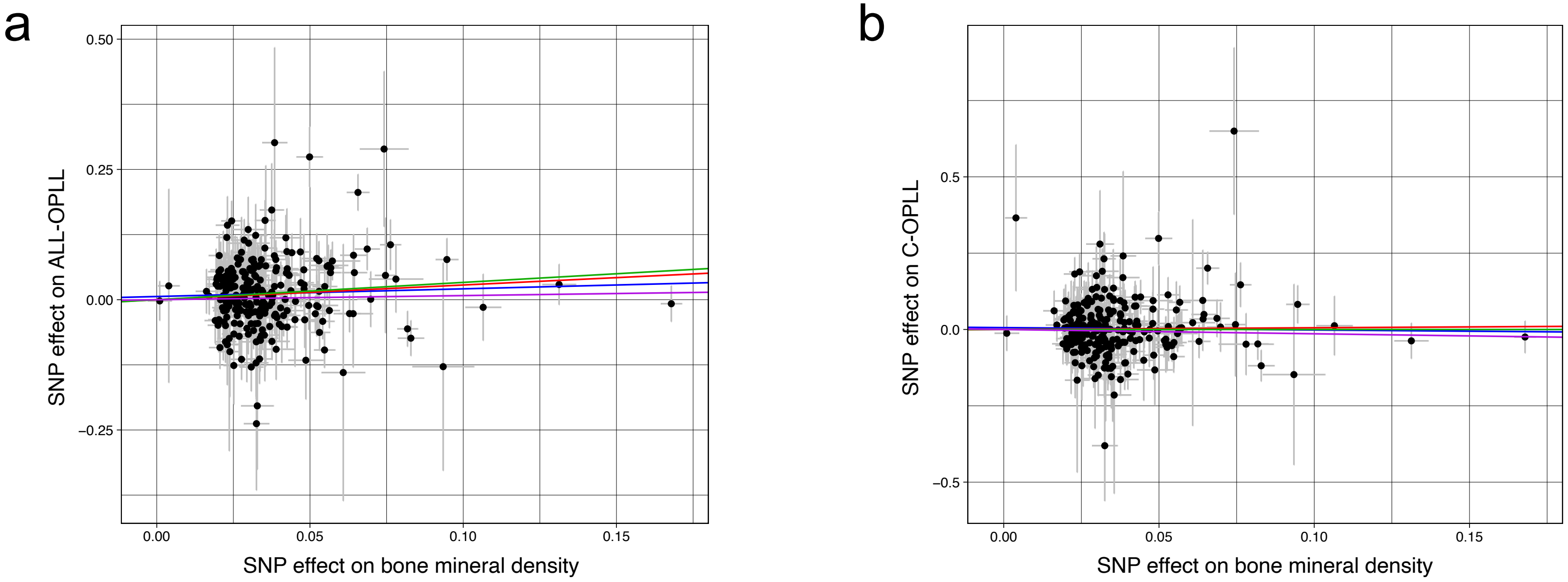


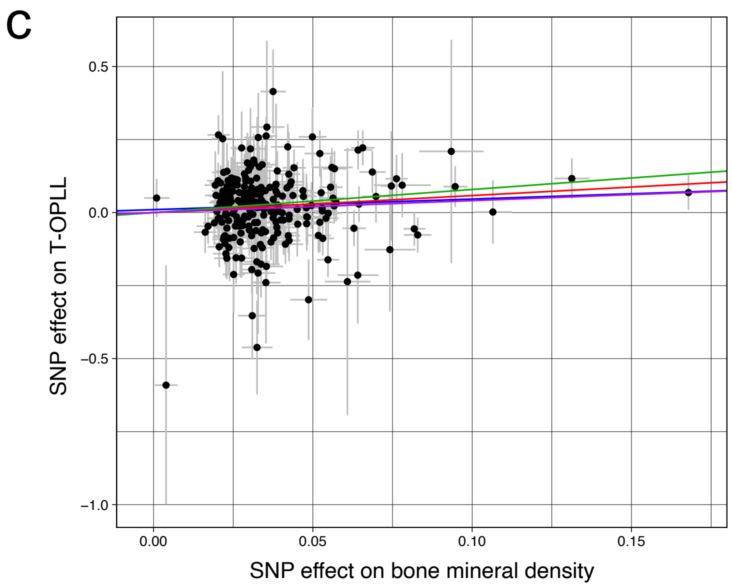


**Supplementary Fig. 13 | Scatter plots for the Mendelian randomization of the causal effect of bone mineral density on OPLL**

(a) OPLL. (b) cervical OPLL. (c) thoracic OPLL. Each dot represents the bone mineral density-associated SNP plotted along with effect size estimates on bone mineral density (x-axis) and OPLL (y-axis). Error bars represent 95% confidence intervals. The line slopes represent the causal association evaluated by four Mendelian randomization (MR) methods: red, inverse variance weighted (IVW); blue, MR-Egger; green, simple median; purple, weighted median.

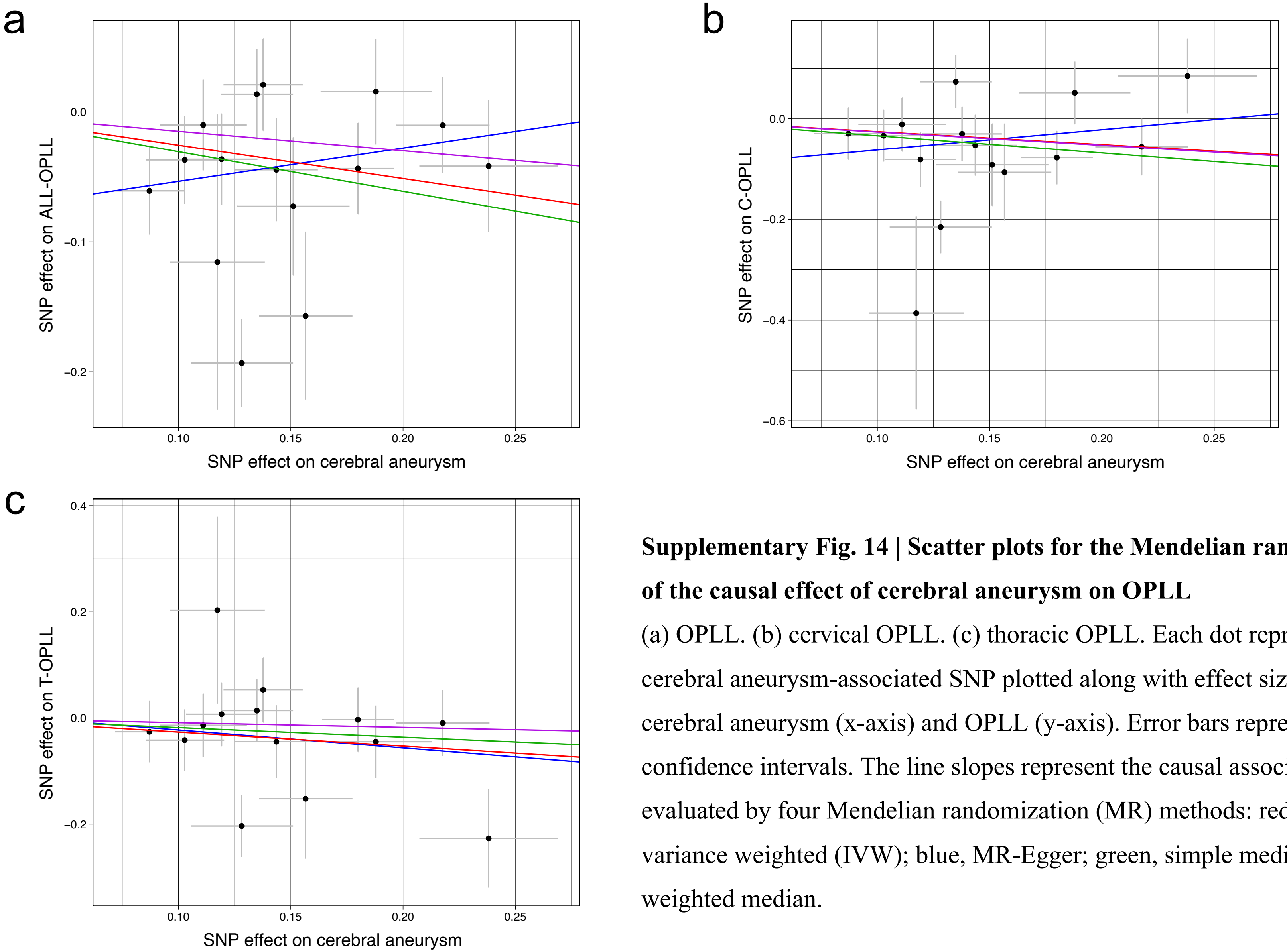


**Supplementary Fig. 14 | Scatter plots for the Mendelian randomization of the causal effect of cerebral aneurysm on OPLL**

(a) OPLL. (b) cervical OPLL. (c) thoracic OPLL. Each dot represents the cerebral aneurysm-associated SNP plotted along with effect size estimates on cerebral aneurysm (x-axis) and OPLL (y-axis). Error bars represent 95% confidence intervals. The line slopes represent the causal association evaluated by four Mendelian randomization (MR) methods: red, inverse variance weighted (IVW); blue, MR-Egger; green, simple median; purple, weighted median.

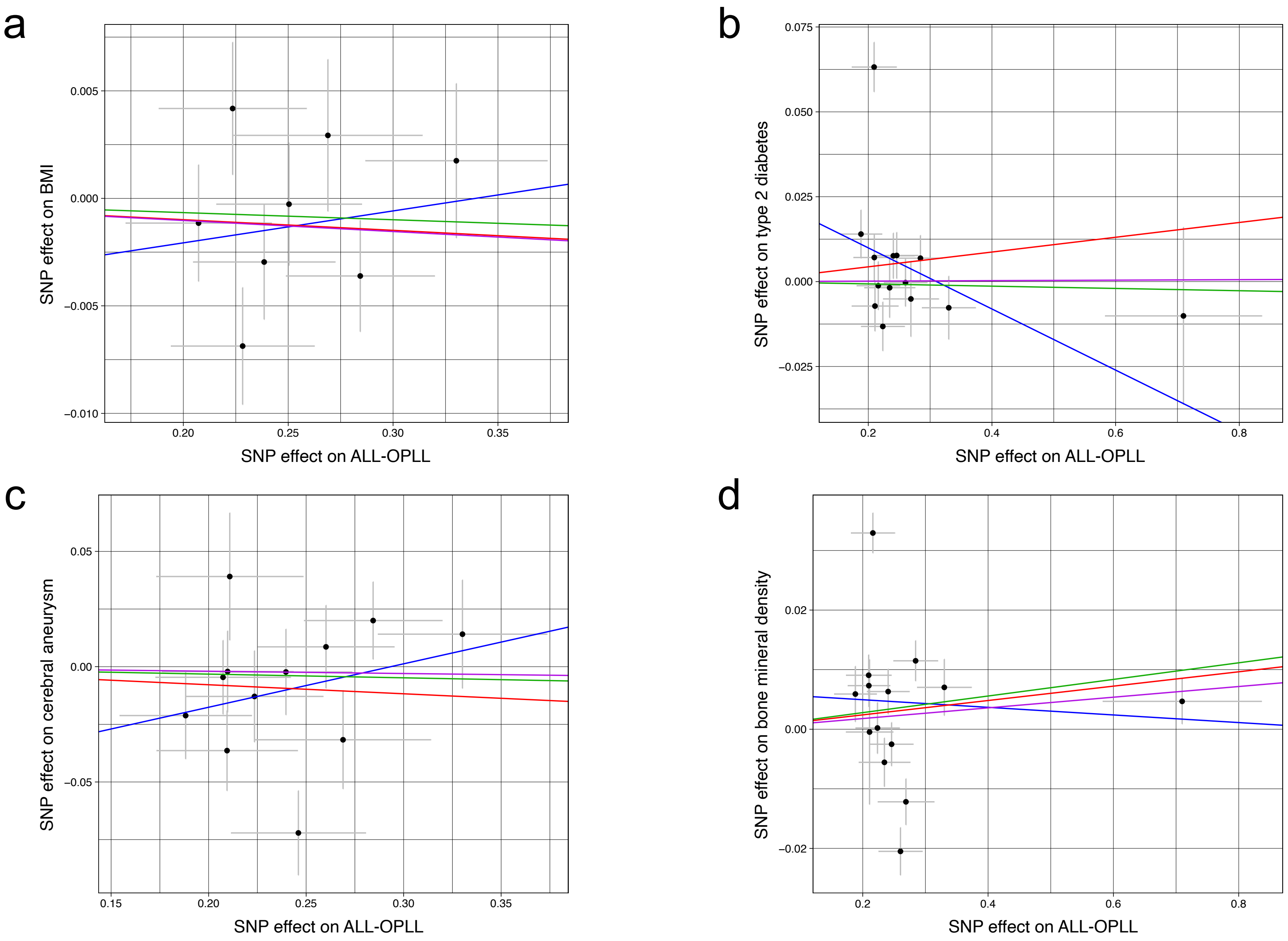


**Supplementary Fig. 15 | See next page for caption**

**Supplementary Fig. 15 | Scatter plots for the Mendelian randomization of the causal effect of OPLL on BMI, type 2 diabetes, cerebral aneurysm, and bone mineral density**

Each dot represents the OPLL-associated SNP plotted along with its effect on OPLL (x-axis) and (a) BMI, (b) type 2 diabetes, (c) cerebral aneurysm, and (d) bone mineral density (y-axis). Error bars represent 95% confidence intervals. The line slopes represent the causal association evaluated by four Mendelian randomization (MR) methods: red, inverse variance weighted (IVW); blue, MR-Egger; green, simple median; purple, weighted median.

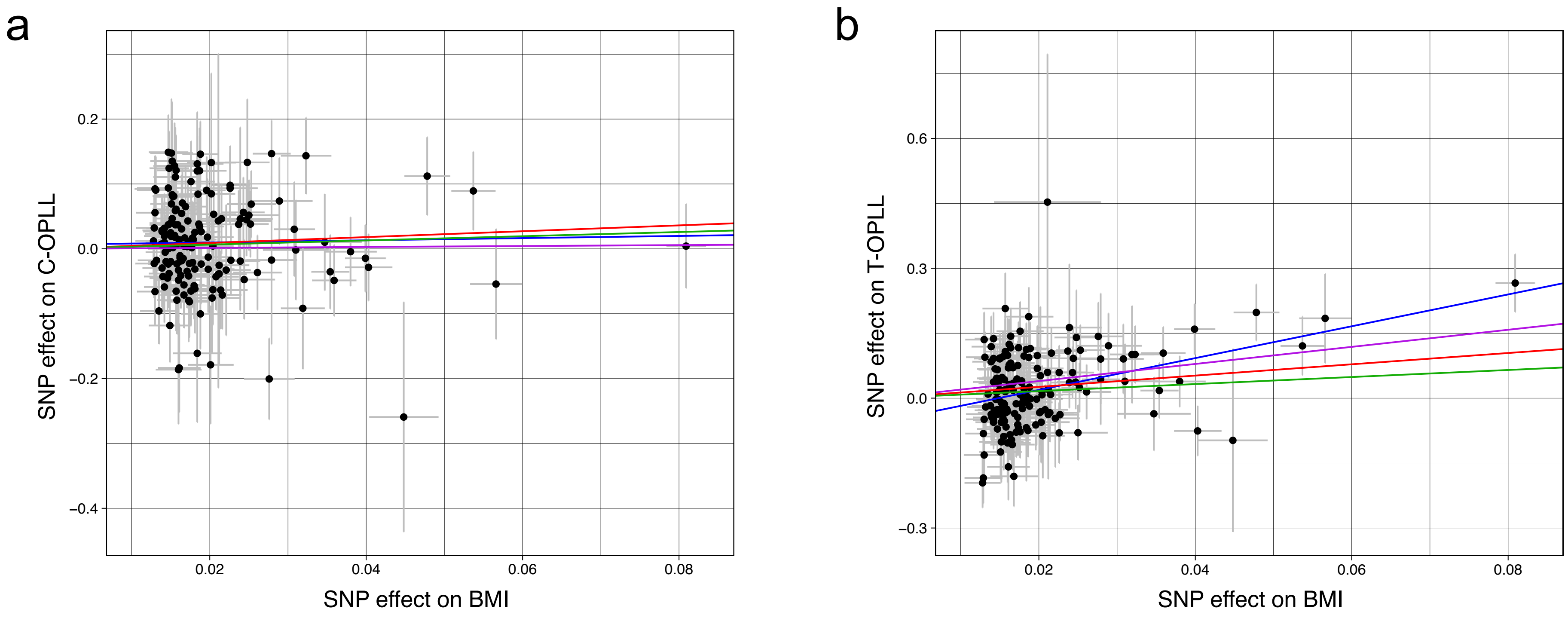


**Supplementary Fig. 16 | Scatter plots for the Mendelian randomization of the causal effect of BMI on OPLL subtypes**

Each dot represents the BMI-associated SNP plotted along with the effect size estimates on BMI (x-axis) and OPLL subtype (y-axis). (a) Cervical OPLL (b) Thoracic OPLL Error bars represent 95% confidence intervals. The line slopes represent the causal association evaluated by four Mendelian randomization (MR) methods: red, inverse variance weighted (IVW); blue, MR-Egger; green, simple median; purple, weighted median.

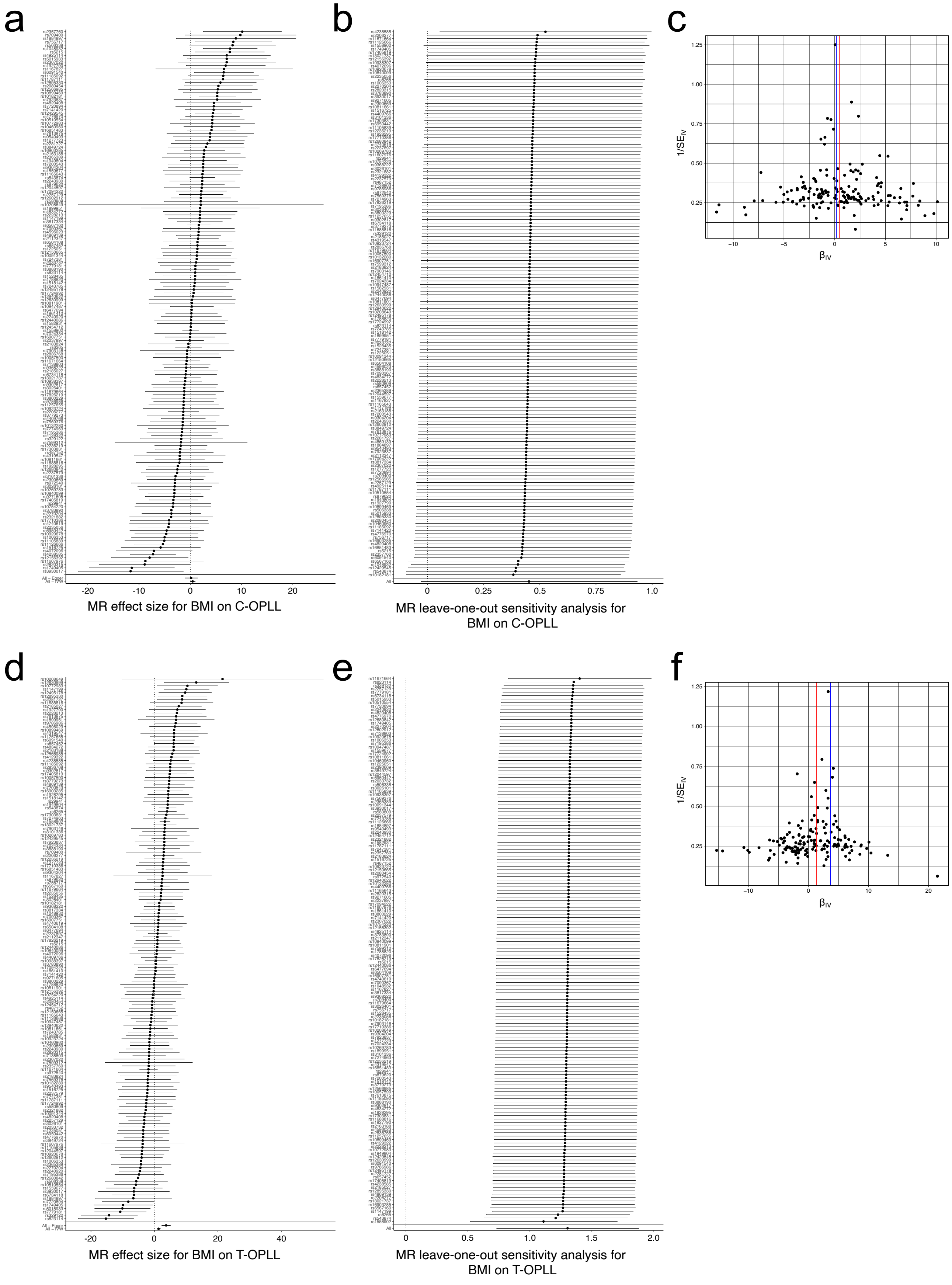


**Supplementary Fig. 17 | See next page for caption**

**Supplementary Fig. 17 | Sensitivity analysis of the Mendelian randomization of BMI causality on OPLL subtypes**

(a), (d) Forest plot. Each black point represents an effect size for BMI on (a) cervical and (d) thoracic OPLL, produced using significant SNPs in BMI GWAS as separate instruments. The black point in the bottom row shows the combined causal estimate using all SNPs together in a single instrument, using two methods of Mendelian randomization (MR): inverse-variance weighted（IVW） and MR-Egger. Horizontal lines are 95% confidence intervals.

(b), (e) Leave-one-out analysis. Each row represents an MR result (IVW) of BMI on (b) cervical OPLL and (e) thoracic OPLL after discarding the SNP listed on the y-axis. The point represents the effect size, and the horizontal line represents 95% confidence intervals.

(c), (f) Funnel plot. On the y-axis, $1/SE_{IV}$ represents the inverse standard error of the estimated causal effect for each of the single SNPs (instrumental variables) ((c) cervical OPLL, (f) thoracic OPLL). On the x-axis, $\beta_{IV}$ represents the effect size of each SNP. Colored lines represent the effect sizes of the different MR analyses: red, IVW; blue, MR-Egger

a

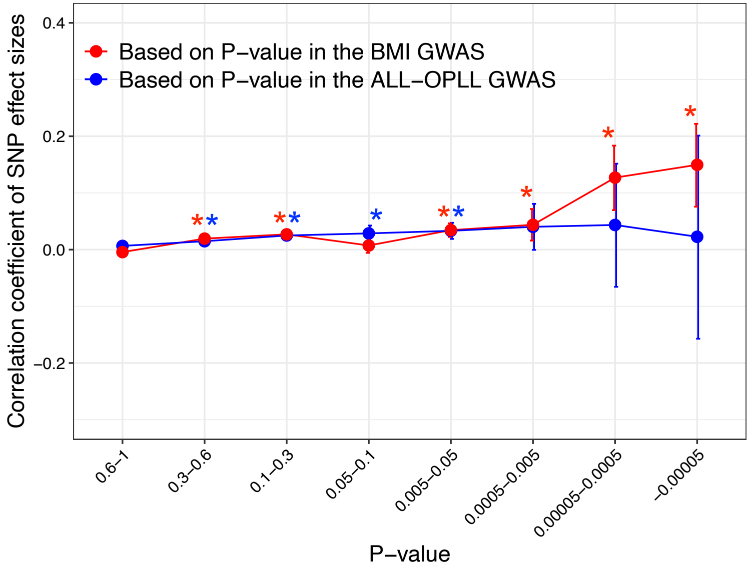


b

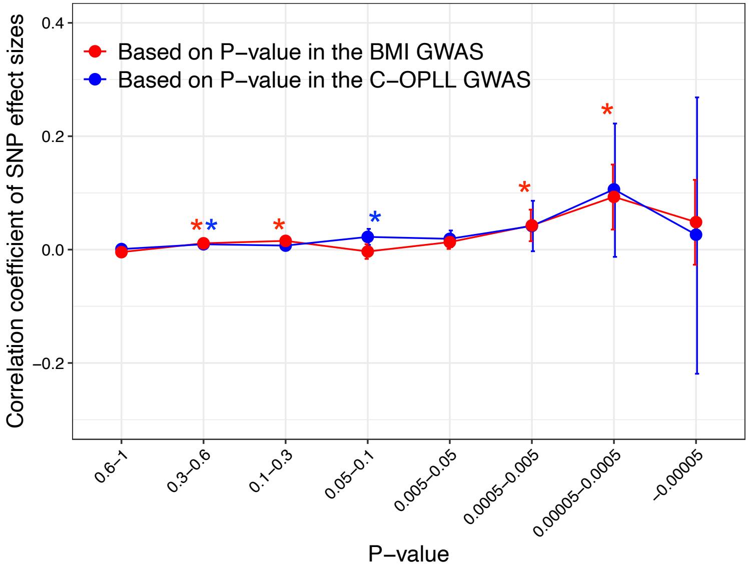


c

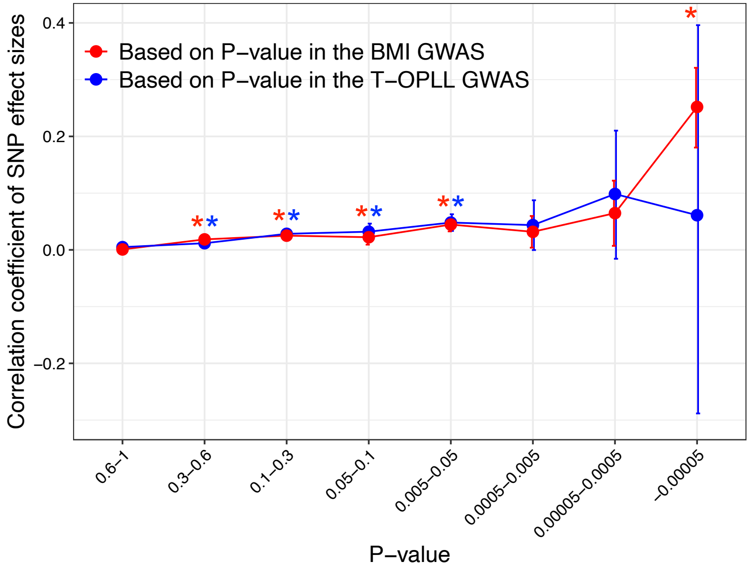


**Supplementary Fig. 18 | Correlation of the effect sizes of the genome-wide SNPs of OPLL and BMI**

(a) ALL-OPLL and BMI, (b) C-OPLL and BMI, (c) T-OPLL and BMI. Correlations were evaluated for sets of SNPs stratified by the thresholds based on the GWAS P-values in each trait. Noted by asterisk is the significant correlation ($P < 0.05/8$). The x-axis shows the P-value of the SNPs, and the y-axis shows the correlation coefficient of the effect size

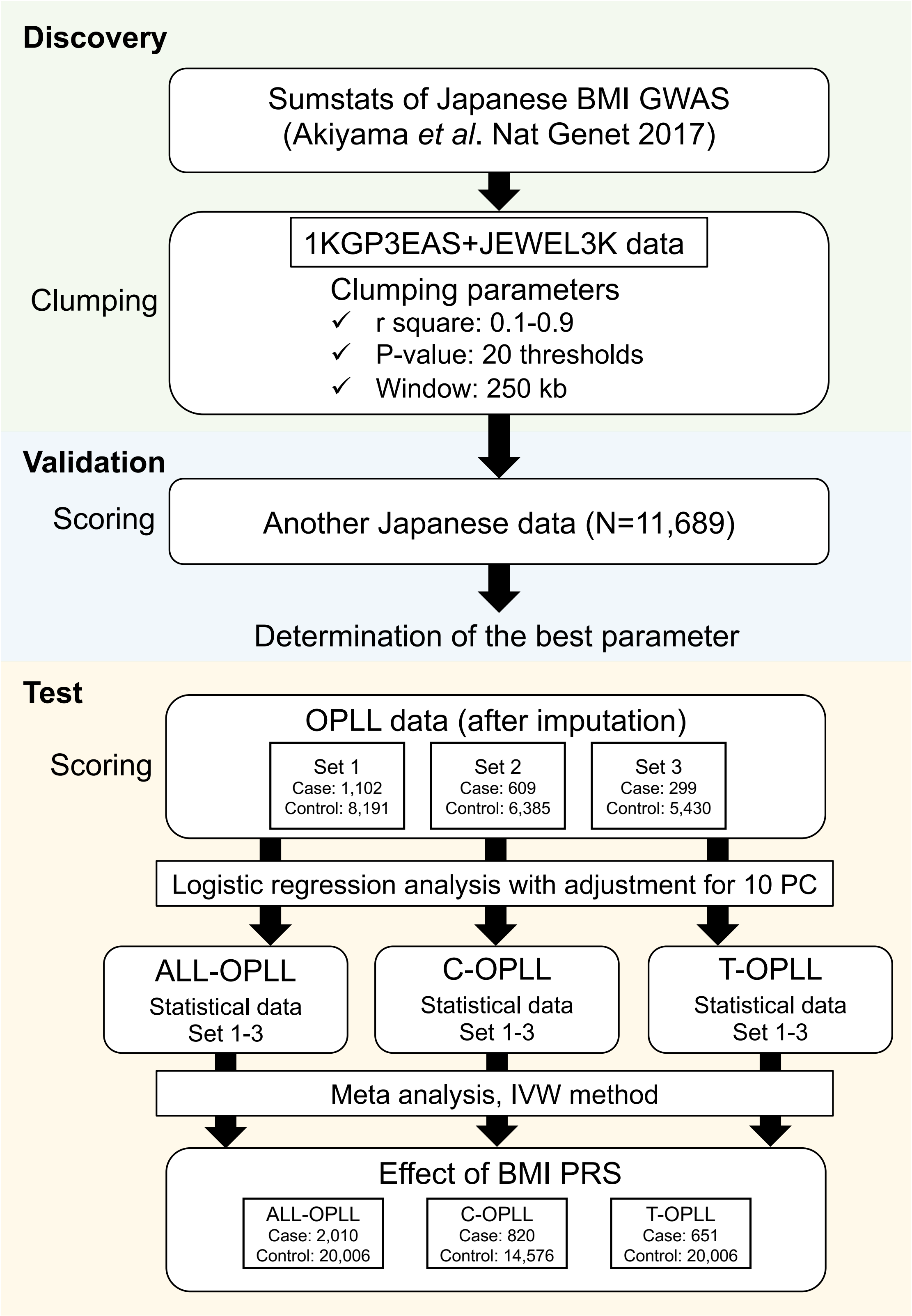


**Supplementary Fig. 19 | BMI polygenic risk score analysis for OPLL**

Overview of the analysis using BMI polygenic risk score for OPLL and its subtypes 1KGP3EAS, the 1000 Genomes Project Phase 3 East Asian; JEWEL_ 3 K, 3,256 in-house Japanese whole-genome sequence data. IVW, inverse-variance weighted.

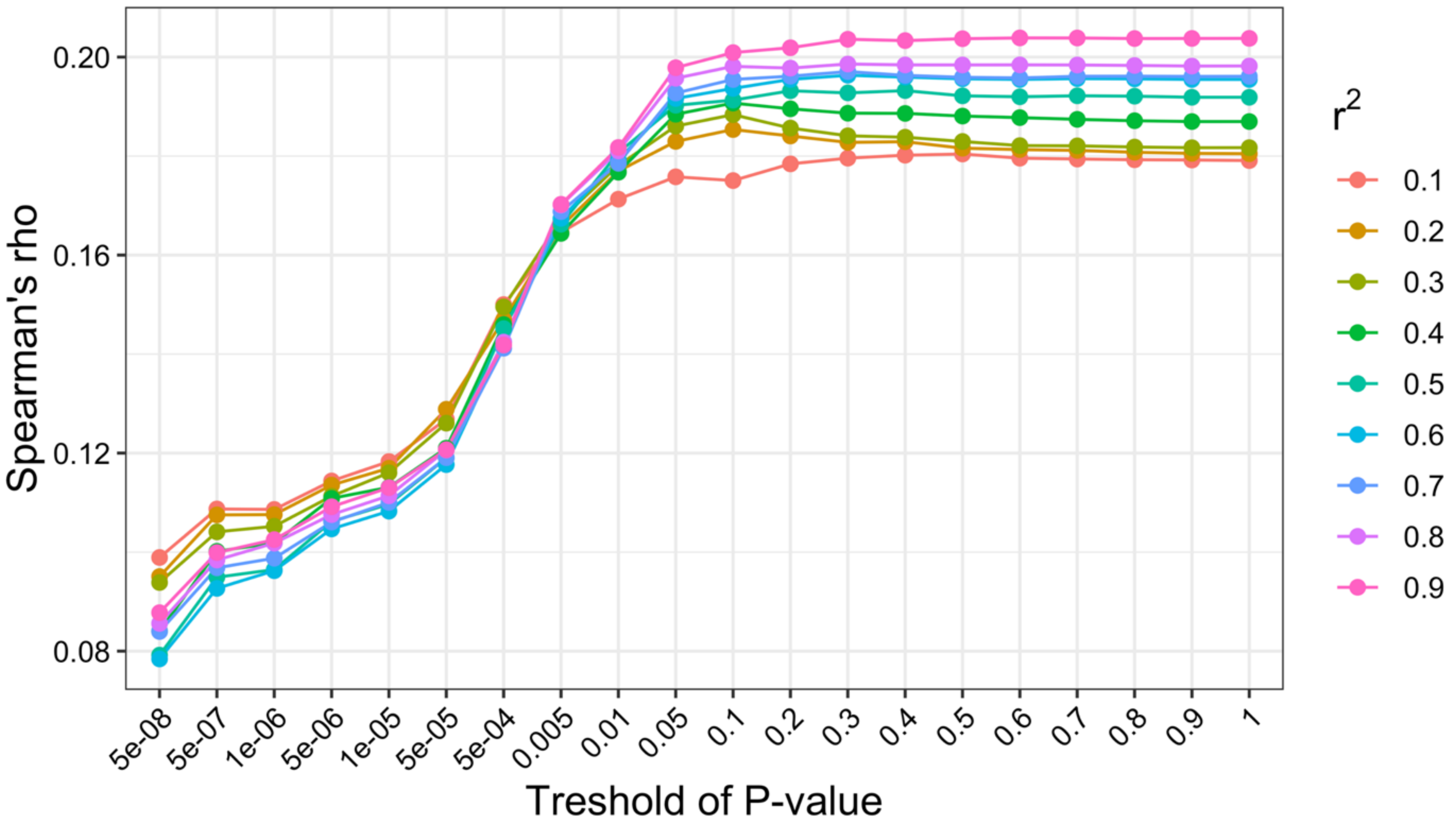


**Supplementary Fig. 20 | Determination of the best parameter for BMI polygenic risk score**

The horizontal line represents the Spearman's rho between the BMI and the BMI polygenic risk score. The vertical line represents the P-value thresholds in clumping. Each color represents the r-square used as a clumping parameter.

**Supplementary Note 1**

**Members of Genetic Study Group of Investigation Committee on Ossification of the Spinal Ligaments**

**Takashi Tsuji, Takeshi Miyamoto, Kazuhiro Chiba, Morio Matsumoto, Yoshiaki Toyama**

Department of Orthopaedic Surgery, School of Medicine, Keio University, Tokyo, Japan.

**Hiroyuki Inose, Toshitaka Yoshii, Shigenori Kawabata, Atsushi Okawa**

Department of Orthopaedic Surgery, Tokyo Medical and Dental University, Tokyo, Japan.

**Masashi Yamazaki, Koda Masao**

Department of Orthopaedic Surgery, Faculty of Medicine, University of Tsukuba, Tsukuba, Japan

**Yoshinao Koike, Masahiko Takahata, Tsutomu Endo**

Department of Orthopedic Surgery, Hokkaido University Graduate School of Medicine, Sapporo, Japan.

**Shiro Imagama, Kazuyoshi Kobayashi, Hiroaki Nakashima, Kei Ando**

Department of Orthopedics, Nagoya University Graduate School of Medicine, Nagoya, Japan.

**Takashi Kaito, Masafumi Kashii**

Department of Orthopaedic Surgery, Osaka University Graduate School of Medicine, Osaka, Japan.

**Satoshi Kato**

Department of Orthopaedic Surgery, Graduate School of Medical Science, Kanazawa University, Kanazawa, Japan.

**Yoshiharu Kawaguchi**

Department of Orthopaedic Surgery, Toyama University, Toyama, Japan.

**Hiroaki Sakai**

Department of Orthopaedic Surgery, Spinal Injuries Center, Iizuka, Japan.

**Shigeo Shindo**

Department of Orthopedics, Kudanzaka Hospital, Tokyo, Japan.

**Yuki Taniguchi**

Department of Orthopaedic Surgery, Faculty of Medicine, The University of Tokyo, Tokyo, Japan.

**Kazuhiro Takeuchi**

Department of Orthopaedic Surgery, National Okayama Medical Center, Okayama, Japan.
**Shingo Maeda, Kawamura Ichiro**
Department of Medical Joint Materials, Graduate School of Medical and Dental Sciences, Kagoshima University, Kagoshima, Japan.
**Hideaki Nakajima, Hisatoshi Baba, Kenzo Uchida**
Department of Orthopaedics and Rehabilitation Medicine, Faculty of Medical Sciences, University of Fukui, Fukui, Japan.
**Kanji Mori**
Department of Orthopaedic Surgery, Shiga University of Medical Science, Otsu, Japan.
**Atsushi Seichi, Atsushi Kimura**
Department of Orthopedics, Jichi Medical University, Shimotsuke, Japan.
**Shunsuke Fujibayashi**
Department of Orthopaedic Surgery, Graduate School of Medicine, Kyoto University, Kyoto, Japan.
**Tsukasa Kanchiku**
Department of Orthopedic Surgery, Yamaguchi University Graduate School of Medicine, Ube, Japan.
**Kei Watanabe**
Department of Orthopaedic Surgery, Niigata University Medical and Dental General Hospital, Niigata, Japan.
**Toshihiro Tanaka**
Department of Orthopaedic Surgery, Hirosaki University Graduate School of Medicine, Hirosaki, Japan.
**Kazunobu Kida**
Department of Orthopaedic Surgery, Kochi Medical School, Nankoku, Japan.
**Sho Kobayashi**
Department of Orthopaedic Surgery, Hamamatsu University School of Medicine, Hamamatsu, Japan.
**Masahito Takahashi**
Department of Orthopaedic Surgery, Kyorin University School of Medicine, Tokyo, Japan.
**Kei Yamada**
Department of Orthopaedic Surgery, Kurume University School of Medicine, Kurume, Japan.
**Shiro Ikegawa**

Laboratory for Bone and Joint Diseases, Center for Integrative Medical Sciences, RIKEN, Tokyo, Japan.

**Supplementary Note 2**

**SNP-obesity interaction**

We evaluated the SNP-obesity interaction during the development of OPLL. We first calculated the best-guess genotypes for lead SNPs in 14 significant loci of the OPLL GWAS meta-analysis (ALL-OPLL) using PLINK. We then scored each individual based on the best-guess genotypes of each of the 14 SNPs: risk allele/risk allele = 2, risk allele/non-risk allele = 1, non-risk allele/non-risk allele = 0. Furthermore, we scored each individual according to the WHO classification of obesity: BMI $< 18.5$ (underweight) scored 0; $18.5 \leq$ BMI $< 25.0$ (normal range) scored 1; $25.0 \leq$ BMI $< 30.0$ (overweight) scored 2; $30.0 \leq$ BMI $<35.0$ (obese class 1) scored 3; $35.0 \leq$ BMI $< 40.0$ (obese class 2) scored 4; and $40.0 \leq$ BMI (Obese class 3) scored 5 (World Health Organization, 2000). We measured the association between the interaction term (SNP*obesity score) and OPLL using logistic regression with principal components 1-10 obtained by principal component analysis with SmartPCA (Patterson et al., 2006) as covariates.

We found no significant association between the development of OPLL and SNP-obesity interaction.
